# Who seeks care, and what gets measured? Understanding the distinct mechanisms behind visit and observation processes in multi-center electronic health records

**DOI:** 10.64898/2026.08.12.26360236

**Authors:** Cheng-Han Yang, Maxwell Salvatore, Haidong Lu, Zihan Zhu, Peter Tennant, Xu Shi, Lucila Ohno-Machado, Rohan Khera, Cary Gross, Fan Li, Bhramar Mukherjee

**Affiliations:** Department of Biostatistics, Yale School of Public Health, New Haven, CT 06510, USA; Department of Biostatistics, Epidemiology and Informatics, Perelman School of Medicine, University of Pennsylvania, Philadelphia, PA 19104, USA; Department of Genetics, Perelman School of Medicine, University of Pennsylvania, Philadelphia, PA 19104, USA; Section of General Internal Medicine, Department of Internal Medicine, Yale School of Medicine, New Haven, CT 06511, USA; Department of Chronic Disease Epidemiology, Yale School of Public Health, New Haven, CT 06510, USA; Department of Biostatistics, School of Public Health, University of Michigan, Ann Arbor, MI 48109, USA; Department of Biomedical Informatics and Data Science, Yale School of Medicine, New Haven, CT 06510, USA; Section of Cardiovascular Medicine, Department of Internal Medicine, Yale School of Medicine, New Haven, CT 06511, USA; Department of Statistics and Data Science, Yale University, New Haven, CT 06511, USA

## Abstract

Electronic health record (EHR)-linked cohorts support association, prediction, and causal studies using longitudinally measured markers of health. However, a lab biomarker measurement is recorded only when a patient first has a medical encounter (visit process) and, a clinician orders the corresponding test and the patient follows through (observation process). These two stages may induce informative presence (IP) and informative observation (IO), respectively. Yet their drivers remain largely uncharacterized, despite evidence that understanding this recording mechanism is essential for selecting appropriate strategies for downstream analysis that treat these markers as longitudinally measured outcomes. We characterize this two-stage recording hierarchy using a stochastic recurrent-event model for the outpatient visit process and a visit-process-weighted generalized estimating equation model for biomarker recording conditional on an outpatient visit. We characterize descriptors of both processes in three EHR-linked cohorts in the US (All of Us [AoU], n=599,423; Yale New Haven Health System [YNHHS], n=319,666; Michigan Genomics Initiative [MGI], n=82,372), reporting descriptive statistics for longitudinal visits and for a panel of 68 lab biomarkers commonly measured in EHRs. We conduct detailed model-based analyses of ten biomarkers spanning multiple domains: routine monitoring, general laboratory assessment, and symptom-triggered testing. These include glucose, hemoglobin A1c [HbA1c], creatinine, hemoglobin [Hgb], white blood cell count [WBC], low-density lipoprotein [LDL] and high-density lipoprotein [HDL] cholesterol, triglycerides, C-reactive protein [CRP], and thyroid-stimulating hormone [TSH]. Across the three cohorts, the median number of outpatient visits ranged from 1.7 to 6.1 per year over a median follow-up of 4.4 to 7.2 years. Among patients with at least one recorded measurement, the median within-person proportion of visits containing a given biomarker ranged from 0.4% to 19.5%, demonstrating that more frequent visits did not necessarily translate into greater per-visit biomarker capture. In the visit-process models, chronic disease burden, and a recent history of outpatient visits were consistently associated with higher visit rates across all three cohorts whereas associations with race, ethnicity, and neighborhood-level income varied across cohorts. In per-visit observation models, the association of covariates depended on the biomarker under consideration; for example, prior cancer diagnosis was associated with more frequent measurement of blood counts but with less frequent measurement of lipids. These findings provide a deeper understanding of how to model who seeks care and what is measured as two distinct recording processes in EHR. Our empirical findings show that the descriptors of these processes vary across cohorts and biomarkers, providing guidance on how to construct these models for downstream longitudinal analyses with irregular EHR visits.

## Introduction

Electronic health records (EHRs) and biobanks have become foundational resources for genome-wide, phenome-wide and lab-wide association studies, disease risk prediction, and causal effect estimation^1–4^. These resources capture rich longitudinal clinical measurements for large numbers of patients under routine clinical practice. Unlike a designed cohort study with prespecified visits and follow-up schedules, an EHR database records a lab value only as a byproduct of routine clinical care. Who seeks care and what is measured at each encounter depend on patient, clinical, and healthcare-system factors, including barriers to attending visits and completing recommended tests, rather than an investigator-driven sampling design. Associations estimated from EHR data may therefore reflect the underlying disease process, patient characteristics, barriers to care, and the clinical processes determining who is seen and what is measured. Prior work has described this stochastic recording structure as informative presence^4–7^ or an informative visit process^8^. Traditional longitudinal methods with these biomarkers as outcomes (regressed against risk factors), often assume non-informative visit times, an assumption that can fail when testing responds to health indicators or symptoms.

The first step in the recording process is having an encounter or a visit. It is well characterized that inclusion in an EHR and the number of recorded encounters reflect patient sociodemographic characteristics underlying morbidity and clinical indication^9,10^, so that patients observed in an EHR represent a selected and generally sicker subset of the source population^5^. Attendance at scheduled visits is also shaped by patient, healthcare-system, and neighborhood factors, including unmet social needs, insurance, language, and socioeconomic disadvantages^11^. Outpatient visit rates are reported to be higher among women and at the extreme ranges of age distribution in national ambulatory data^12^. In UK Biobank and All of Us, conditions linked to greater healthcare contact showed elevated short-term hazards of incident dementia that attenuated over follow-up, consistent with enhanced detection through referral and healthcare utilization^13^. The second step concerns what is measured once a patient is seen. Conditional on a visit, whether a test is ordered and ultimately recorded can vary by patient characteristics, clinician practices, and clinical indications. Some tests are used for routine monitoring, others primarily to assess acute illness, and others in both contexts. For example, racial and ethnic minority patients are reported to receive advanced imaging tests less often than White patients across care settings and modalities^14^. Documentation of diagnoses and family history also varies by sex, race, ethnicity and language preference, with consequences for the clinical algorithms that rely on these records^15,16^. These measurement gaps accumulate, so that minority patients are evaluated over more visits before the same diagnosis is reached^17^. Differential capture of comorbidities between Medicare Advantage and Traditional Medicare beneficiaries produced large, spurious differences in risk-adjusted outcomes^18^. In statistical terms, the **visit process** generates irregular outpatient visits and may induce bias in association estimates from a linear mixed model from informative presence (IP). Conditional on a visit, biomarker-specific **observation processes** then determine whether a biomarker is recorded and may induce bias in association estimates from informative observation (IO).

Advanced methods developed to address IP propagating through the visit process^19–23^, such as inverse-intensity weighting and joint modeling for irregular visits, leave IO unaddressed. These methods implicitly assume that, once a patient visits, the biomarker of interest is recorded, creating a complete data grid. For example, consider a hypothetical patient with diabetes and no other health conditions who had 25 encounters over five years of follow-up. Hemoglobin A1c (HbA1c) was recorded every six months to represent routine diabetes monitoring, while prostate-specific antigen (PSA) was recorded annually, coinciding with every other HbA1c measurement, to represent periodic screening. Glucose was recorded at each HbA1c encounter and additional encounters, whereas white blood cell count (WBC) was measured less systematically. Glucose, HbA1c, WBC and PSA were recorded at 64%, 44%, 32% and 24% of encounters, respectively (Figure 1a). Moreover, both visit and observation processes (Figure 1b) vary across patients, such that patients with more visits do not necessarily contribute more information for a given biomarker. For example, Patient 1 (Figure 1c) had WBC recorded at only four of 13 visits (31%), whereas Patient 2 had WBC recorded at all six visits (100%). Methods that adjust only for the visit process assign weights to patients according to how often they are seen rather than how often the biomarker of interest is measured. As described in Figure 1c, when WBC is the biomarker of interest, such visit-based weighting can misallocate weights across patients and may perform worse than an unweighted standard linear mixed model^25^.

**Figure 1.**
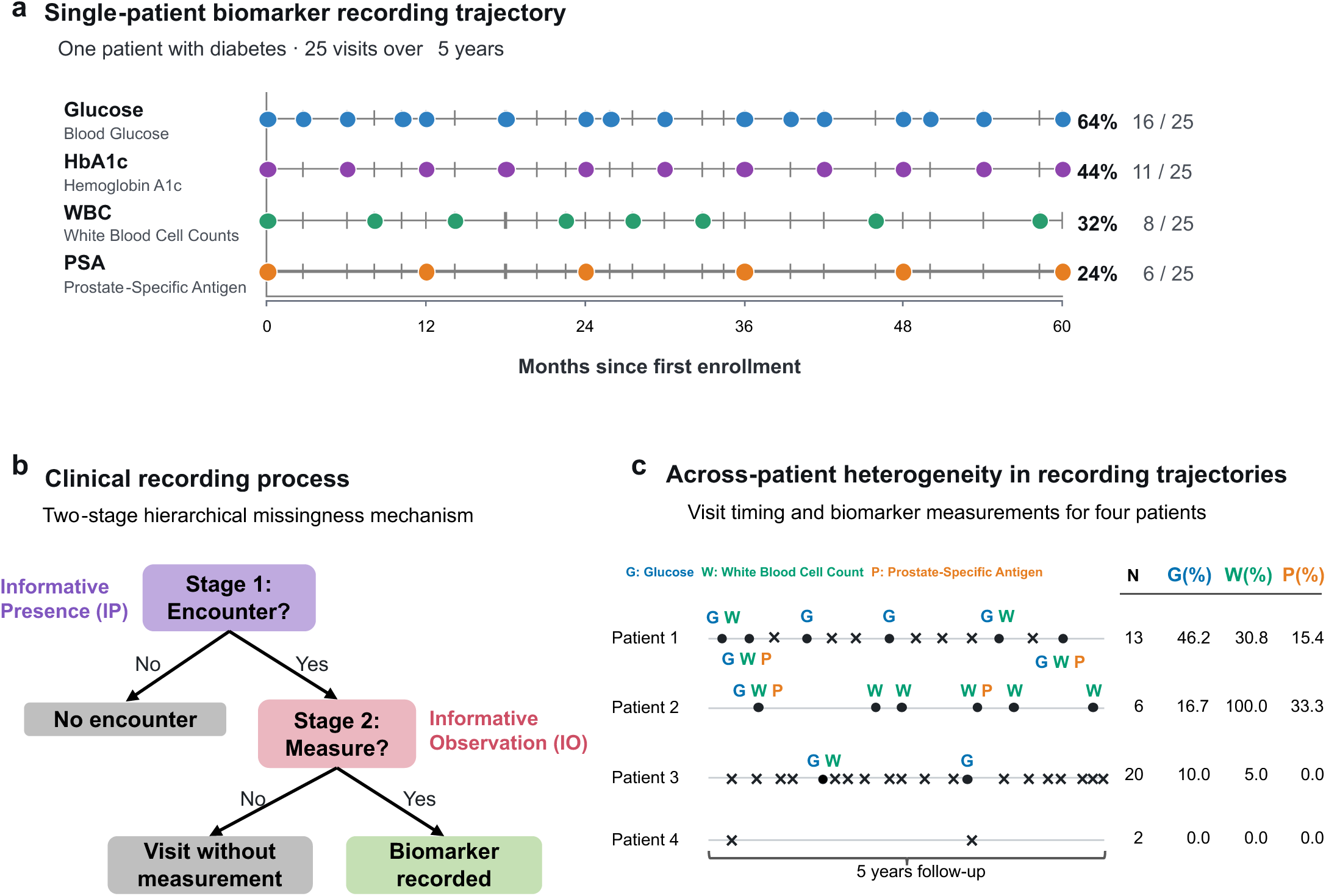
Clinical recording of EHR biomarkers as a two-stage process. a, Recording trajectory for an illustrative patient with diabetes over five years (25 outpatient visits). Hemoglobin A1c (HbA1c) was recorded every six months to represent routine diabetes monitoring, and prostate-specific antigen (PSA) was recorded annually, coinciding with every other HbA1c measurement, to represent periodic screening. Glucose was recorded at each visit with an HbA1c measurement and at additional visits, whereas white blood cell count (WBC) was recorded on a more ad hoc basis. Glucose, HbA1c, WBC and PSA were recorded at 64%, 44%, 32% and 24% of visits, respectively, showing that a visit is necessary but not sufficient for a biomarker to be recorded. b, The recording process as a two-stage hierarchical missingness mechanism: Stage 1 (visit) determines healthcare contact, and Stage 2 (measurement), conditional on a visit, determines whether a biomarker is recorded. Either stage can be informative for the biomarker trajectory, yielding informative presence (IP) and informative observation (IO). c, Visit and measurement trajectories for four representative patients. Crosses denote visits without measurement; filled circles denote visits with glucose (G), WBC (W) or PSA (P) recorded. Visit timing and biomarker-specific measurement frequency vary substantially across patients.

Heterogeneity in visit and measurement frequency does not, by itself, distort a given association target. As in the classical missing-data literature, whether it biases the target association depends on what drives recording (Figure 2). Measured demographics (such as age, sex, race, and ethnicity), healthcare access (such as insurance, income, and rural or urban residence) and comorbidities (such as diabetes, hypertension, and cancer) all shape how often a patient is seen and which tests are ordered. When recording depends only on such observed characteristics, the recorded data are missing at random (MAR), and the target association may be recovered by appropriately adjusting for them. A more challenging scenario is a missing-not-at-random (MNAR) mechanism, in which recording depends on the unrecorded value itself (the orange paths in Figure 2): symptoms suggestive of anemia may prompt a blood-count panel, whereas suspected thyroid disease may prompt a thyroid panel. Lab values may therefore be more likely to be abnormal when a test is ordered, and adjustment for observed covariates alone may not remove the resulting bias. Observation processes are also likely to differ across biomarkers; for example, a screening test recommended after age 50 such as mammography or colonoscopy may plausibly be MAR given patient age, whereas symptom-triggered diagnostic tests (TSH for example) are more likely to be MNAR. Recent methodological work suggests that accounting for both the visit and observation processes is critical for valid longitudinal association analyses^24,26,25^.

**Figure 2.**
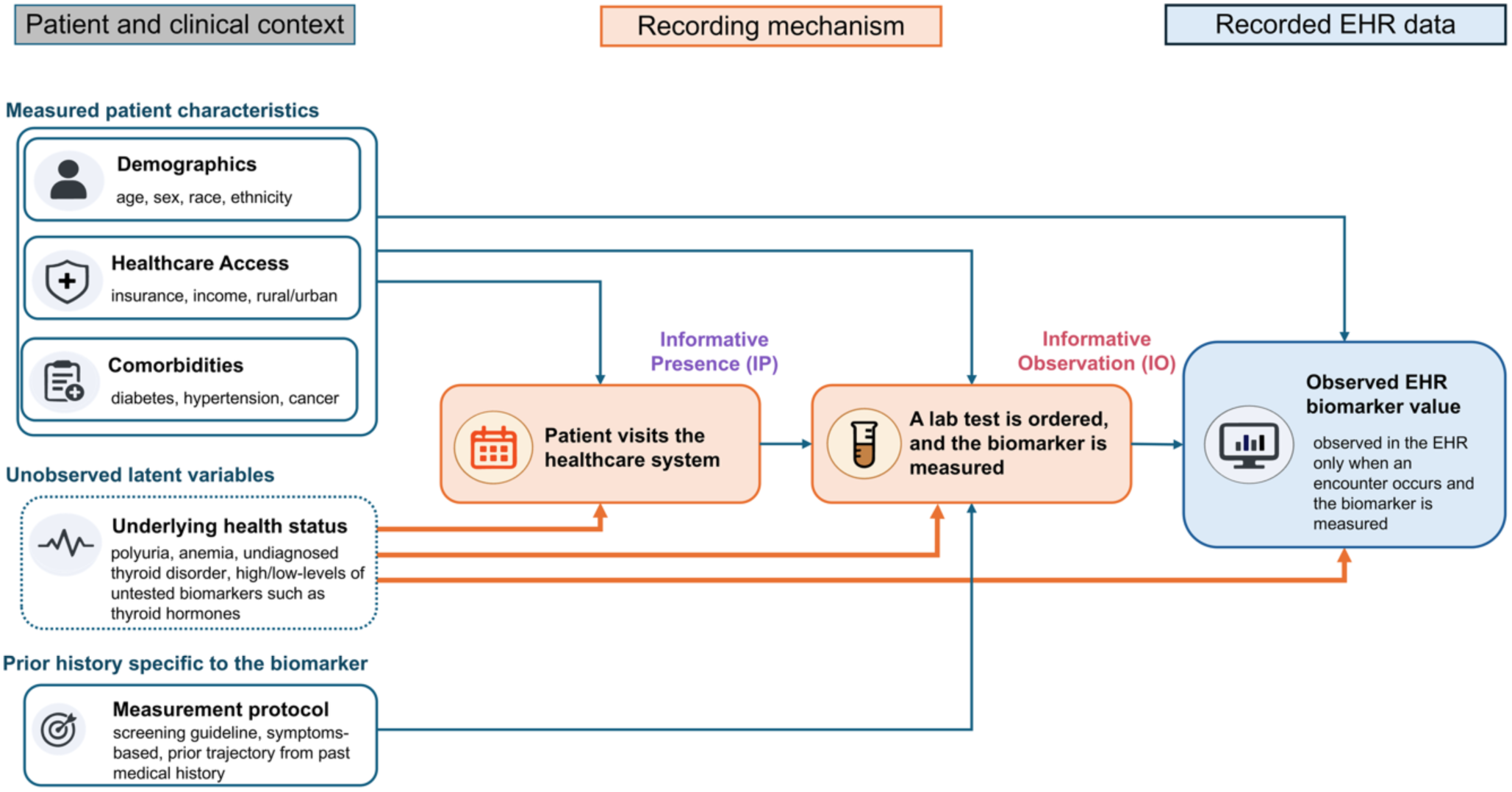
Directed acyclic graph illustrating how the visit and observation processes can bias EHR biomarker analyses. Schematic showing how unobserved heterogeneity propagates through the two-stage clinical recording process to bias analyses of EHR biomarker data. An unobserved latent factor representing underlying health status jointly drives outpatient visits, biomarker recording at a visit, and the biomarker value itself. A biomarker value is therefore observed only through a sequence of selections, each linked to the same latent factor: this opens two distinct pathways through which latent health status confounds the relationship between observed covariates and the biomarker. Adjusting for the visit process closes one pathway but leaves the other open, so estimates of covariate effects on the biomarker remain biased; consistent estimation requires modelling both stages jointly.

This paper has three aims. The *first* is to raise awareness around this complex recording structure inherent in EHR data through a descriptive empirical analysis of patient visits and occurrence of lab tests for 68 commonly measured biomarkers (used in LabWAS^4^). The *second* is to identify which patient characteristics are associated with how often a patient is seen. The *third* is to identify which patient characteristics are associated with whether a biomarker is recorded, and to show that these characteristics are not the same as those associated with visiting.

We selected three EHR-linked US cohorts with contrasting recruitment and data-capture structures. All of Us (AoU)^27^ links records from multiple unaffiliated healthcare settings and oversamples populations historically underrepresented in biomedical research; Yale New Haven Health System (YNHHS)^28^ is a regional Epic-based integrated health system serving a racially diverse Connecticut population; and the Michigan Genomics Initiative (MGI)^29^ is a single-center biobank with perioperative-enriched recruitment.

For the first aim, we characterize recording patterns in these three cohorts: who is seen, which biomarkers are measured, the fraction of patients measured, and the proportion of visits containing a lab result. We profile the commonly measured 68-biomarker panel descriptively and restrict the model-based analysis to ten biomarkers, spanning different clinical ordering logics. For example, lipid panels^30^ and HbA1c^31^, follow guideline-recommended intervals for patients on statins or with diabetes, whereas others, such as C-reactive protein (CRP)^32^, are used to assess acute inflammation or suspected infection

For the second and third aims, we characterize the visit and observation processes, respectively, through models. Let *N_i_*(*t*) denote the cumulative number of outpatient visit-days for patient *i* by time *t*, 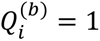 indicate whether patient *i* is ever measured for biomarker *b* during their entire follow-up, and 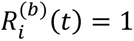 indicate that biomarker *b* is recorded at a visit at time *t*. We model *N_i_*(*t*) using a recurrent-event model for outpatient visits^33^, 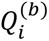 using logistic regression for the probability of at least one measurement over follow-up, and the per-visit observation process 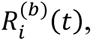 conditional on a visit, using a generalized estimating equation (GEE)^34^ with weights derived from the visit process.

Prior large-scale EHR analyses, including lab-wide association studies^4,35^, typically reduce repeated measurements of each biomarker to a single value (often the median) for association analysis, or fit standard linear mixed models. They do not account for differential frequency of visits, follow-up time, or number of measurements. We hope the visit and observation models developed here will guide practitioners in accounting for IP and IO in longitudinal EHR analyses, since they supply key inputs to existing bias-reduction methods^24–26^.

## Results

### Cohort characteristics

The three cohorts differed in their sociodemographic and baseline clinical characteristics (Table 1). AoU (mean age 46.7 yrs) was the most racially and ethnically diverse cohort (16.9% Black, 19.1% Hispanic/Latino), was 59.9% female, and had the lowest prevalence of diabetes (8.1%), hypertension (34.5%), and cancer (14.6%). YNHHS was the youngest cohort (mean age 44.6 yrs), was also racially and ethnically diverse (10.4% Black, 14.3% Hispanic/Latino), had a nearly even sex distribution (50.5% female), and had higher prevalences of diabetes (13.7%), hypertension (39.1%), and cancer (15.5%) than AoU. MGI was the oldest (mean age 56.2 yrs) and least racially and ethnically diverse (6.1% Black, 1.5% Hispanic/Latino) and was 54.0% female. Consistent with its perioperative recruitment, MGI had the highest prevalence of diabetes (21.4%), hypertension (48.8%), and cancer (40.7%). Obesity was common across all three cohorts (40.2% in AoU, 34.8% in YNHHS, and 40.8% in MGI).

**Table 1.**
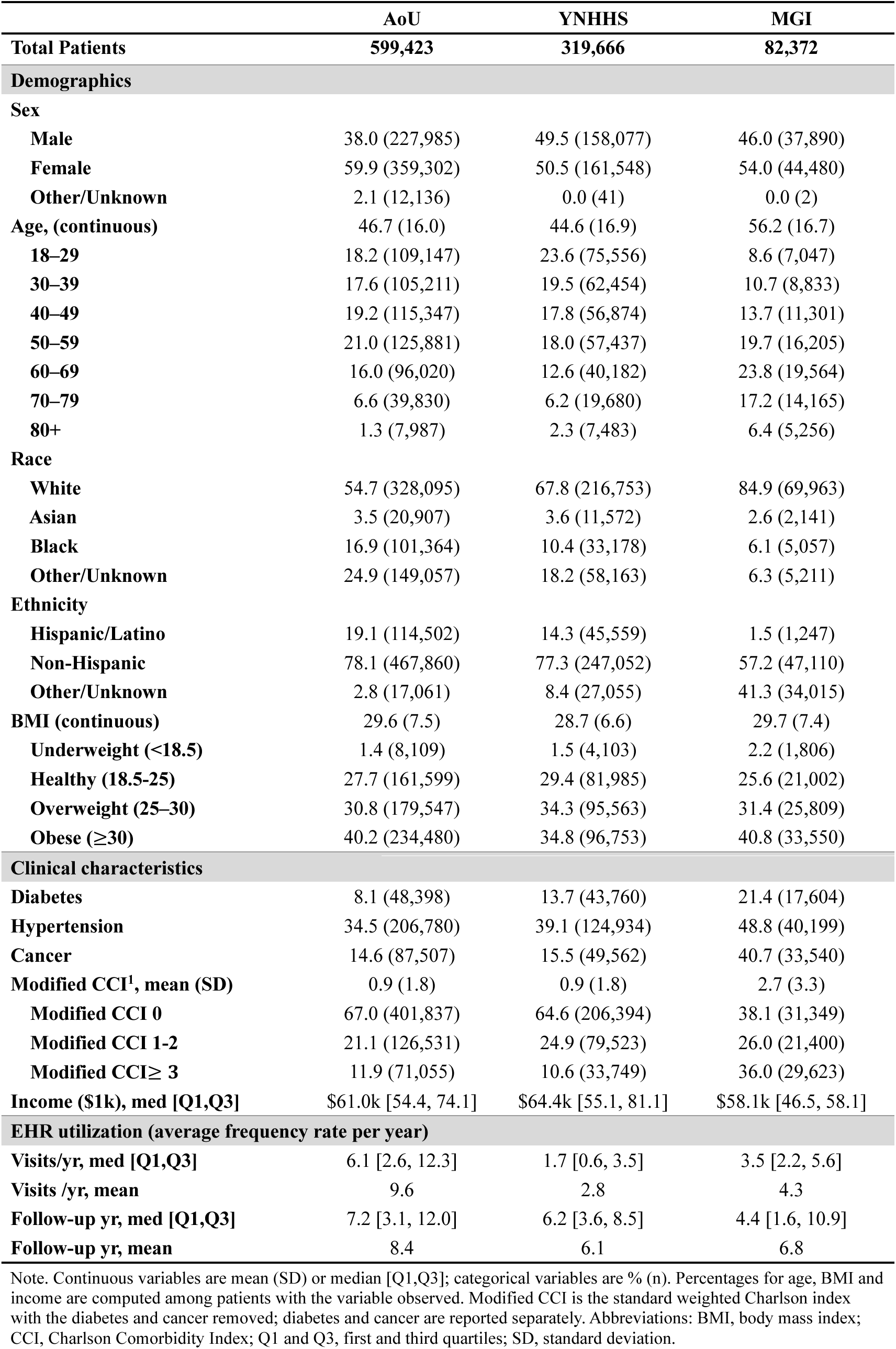
Baseline characteristics of the All of Us (AoU), Yale New Haven Health System (YNHHS), and Michigan Genomics Initiative (MGI). Continuous variables are mean (SD) or median [Q1,Q3]; categorical variables are % (n).

### A. How often patients were seen and which biomarkers were recorded

This is related to our *first descriptive aim* of summarizing the two-step recording across biobanks.

#### Longitudinal visit characteristics

Almost all patients had at least one outpatient visit: 99.5% of AoU, 91.0% of YNHHS, and 99.1% of MGI patients (Table 2). Median follow-up was 7.2, 6.2 and 4.4 years, respectively (Table 1). Among patients with at least one visit, annual visit frequencies differed across cohorts: the median number of outpatient visits was 6.1, 1.9, and 3.5 per year in AoU, YNHHS, and MGI, respectively (Table 2). Total outpatient visits per patient were highly right-skewed (e.g., in AoU, the first, second, and third quartiles were 6, 39, and 137; Table 3).

**Table 2.** Patient characteristics by outpatient visit status in AoU, YNHHS, and MGI cohorts. Continuous variables are reported as mean (SD) or median [Q1,Q3]; categorical variables are reported as % (n). Patients with at least one eligible outpatient visit during follow-up are classified as 1 Visit; those with no qualifying visits are classified as No Visits.

| Characteristic | AoU |  | YNHHS |  | MGI |  |
| --- | --- | --- | --- | --- | --- | --- |
|  | ≥1 Visit | No Visit | ≥1 Visit | No Visit | ≥1 Visit | No Visit |
| **N, % (n)** | 99.5 (596,661) | 0.5 (2,762) | 91.0 (290,935) | 9.0 (28,731) | 99.1 (81,612) | 0.9 (760) |
| **Demographics** |  |  |  |  |  |  |
| **Sex** |  |  |  |  |  |  |
| Female | 59.9 (357,539) | 63.8 (1,763) | 51.1 (148,589) | 45.1 (12,959) | 53.9 (44,012) | 61.6 (468) |
| Male | 38.1 (227,057) | 33.6 (928) | 48.9 (142,320) | 54.8 (15,757) | 46.1 (37,598) | 38.4 (292) |
| **Age, mean (SD)** | 46.7 (16.0) | 42.5 (15.9) | 44.5 (16.8) | 45.1 (18.1) | 56.3 (16.7) | 46.3 (16.3) |
| 18–29 | 18.2 (108,436) | 25.7 (711) | 23.5 (68,513) | 24.5 (7,043) | 8.5 (6,907) | 18.4 (140) |
| 30–39 | 17.5 (104,572) | 23.1 (639) | 19.6 (56,917) | 19.3 (5,537) | 10.6 (8,678) | 20.4 (155) |
| 40–49 | 19.3 (114,875) | 17.1 (472) | 17.9 (51,991) | 17.0 (4,883) | 13.7 (11,140) | 21.2 (161) |
| 50–59 | 21.0 (125,422) | 16.6 (459) | 18.1 (52,776) | 16.2 (4,661) | 19.7 (16,085) | 15.8 (120) |
| 60–69 | 16.0 (95,700) | 11.6 (320) | 12.7 (36,873) | 11.5 (3,309) | 23.8 (19,452) | 14.7 (112) |
| 70–79 | 6.7 (39,700) | 4.7 (130) | 6.0 (17,546) | 7.4 (2,134) | 17.3 (14,112) | 7.0 (53) |
| 80+ | 1.3 (7,956) | 1.1 (31) | 2.2 (6,319) | 4.1 (1,164) | 6.4 (5,237) | 2.5 (19) |
| **Race** |  |  |  |  |  |  |
| White | 54.7 (326,407) | 61.1 (1,688) | 68.9 (200,553) | 56.4 (16,200) | 85.2 (69,512) | 59.3 (451) |
| Asian | 3.5 (20,772) | 4.9 (135) | 3.8 (11,061) | 1.8 (511) | 2.6 (2,090) | 6.7 (51) |
| Black | 17.0 (101,174) | 6.9 (190) | 10.2 (29,606) | 12.4 (3,572) | 6.1 (4,947) | 14.5 (110) |
| Other/Unknown | 24.9 (148,308) | 27.1 (749) | 17.1 (49,715) | 29.4 (8,448) | 6.2 (5,063) | 19.5 (148) |
| **Ethnicity** |  |  |  |  |  |  |
| Hispanic/Latino | 19.1 (113,957) | 19.7 (545) | 14.1 (41,026) | 15.8 (4,533) | 1.5 (1,210) | 4.9 (37) |
| Non-Hispanic | 78.1 (465,714) | 77.7 (2,146) | 78.4 (228,158) | 65.8 (18,894) | 57.1 (46,602) | 66.8 (508) |
| Unknown | 2.8 (16,990) | 2.6 (71) | 7.5 (21,751) | 18.5 (5,304) | 41.4 (33,800) | 28.3 (215) |
| **BMI, mean (SD)** | 29.6 (7.5) | 30.3 (8.1) | 28.7 (6.6) | 28.4 (6.7) | 29.7 (7.4) | 29.8 (7.2) |
| Underweight (<18.5) | 1.4 (8,089) | 1.3 (20) | 1.4 (3,858) | 2.3 (245) | 2.2 (1,799) | 1.1 (7) |
| Healthy (18.5–25) | 27.7 (161,209) | 26.0 (390) | 29.4 (78,634) | 31.0 (3,351) | 25.6 (20,846) | 25.5 (156) |
| Overweight (25–30) | 30.8 (179,092) | 30.4 (455) | 34.4 (92,034) | 32.6 (3,529) | 31.4 (25,616) | 31.6 (193) |
| Obese (≥30) | 40.2 (233,846) | 42.3 (634) | 34.8 (93,060) | 34.1 (3,693) | 40.8 (33,295) | 41.7 (255) |
| **Clinical characteristics** |  |  |  |  |  |  |
| **Diabetes, % (n)** | 8.1 (48,305) | 3.4 (93) | 14.3 (41,568) | 7.6 (2,192) | 21.5 (17,563) | 5.4 (41) |
| **Hypertension, % (n)** | 34.6 (206,277) | 18.2 (503) | 40.8 (118,797) | 21.4 (6,137) | 49.1 (40,103) | 12.6 (96) |
| **Cancer, % (n)** | 14.6 (87,358) | 5.4 (149) | 16.5 (48,046) | 5.3 (1,516) | 41.0 (33,470) | 9.2 (70) |
| **Modified CCI, mean (SD)** | 0.9 (1.8) | 0.3 (0.9) | 0.9 (1.8) | 0.4 (1.1) | 2.8 (3.3) | 0.4 (1.3) |
| Modified CCI 0 | 67.0 (399,539) | 83.2 (2,298) | 63.0 (183,366) | 80.2 (23,028) | 37.6 (30,692) | 86.4 (657) |
| Modified CCI 1–2 | 21.1 (126,147) | 13.9 (384) | 25.8 (75,098) | 15.4 (4,425) | 26.1 (21,332) | 8.9 (68) |
| Modified CCI ≥3 | 11.9 (70,975) | 2.9 (80) | 11.2 (32,471) | 4.4 (1,278) | 36.3 (29,588) | 4.6 (35) |
| **EHR utilization** |  |  |  |  |  |  |
| **Visit/yr, med [Q1,Q3]** | 6.1 [2.6, 12.3] | 0.0 [0.0, 0.0] | 1.9 [0.9, 3.8] | 0.0 [0.0, 0.0] | 3.5 [2.2, 5.6] | 0.0 [0.0, 0.0] |
| **Follow-up yr, med [Q1,Q3]** | 7.2 [3.1, 12.0] | 2.6 [0.9, 5.8] | 6.5 [4.0, 8.7] | 2.3 [0.7, 4.9] | 4.4 [1.6, 10.9] | 0.4 [0.2, 1.1] |

Note: Modified CCI was calculated from the Charlson Comorbidity Index after excluding diabetes and cancer components, which are reported separately.

**Table 3.**
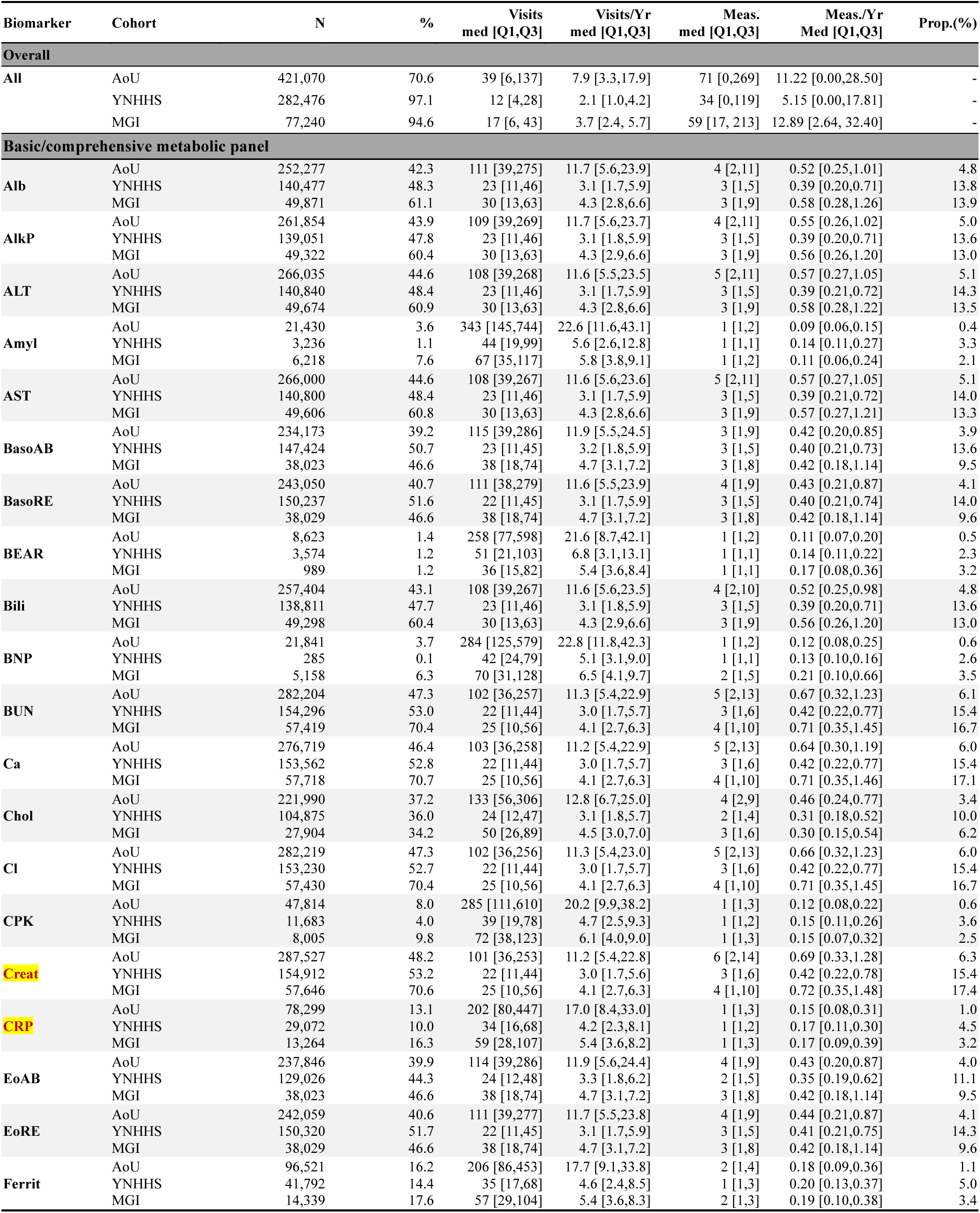
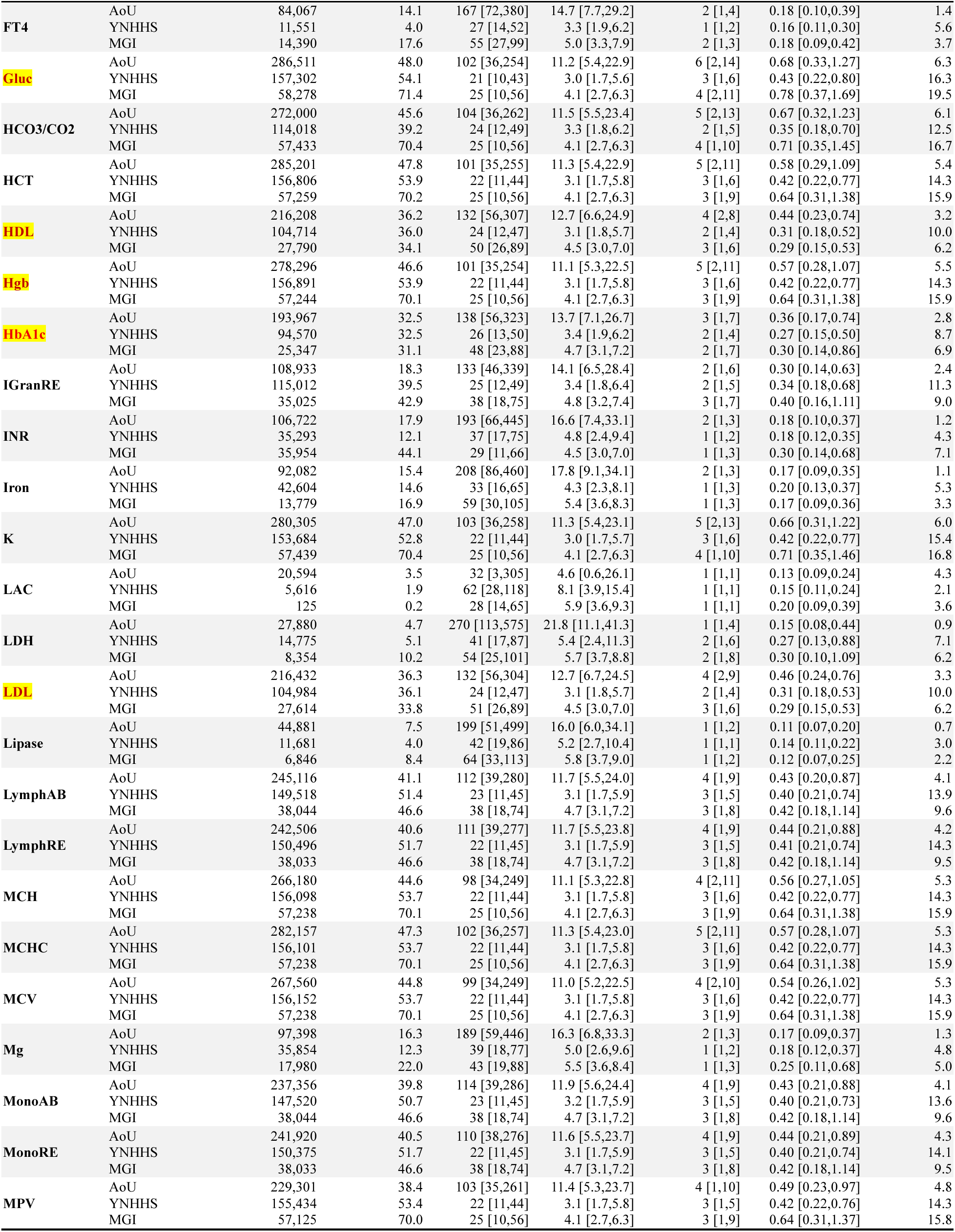
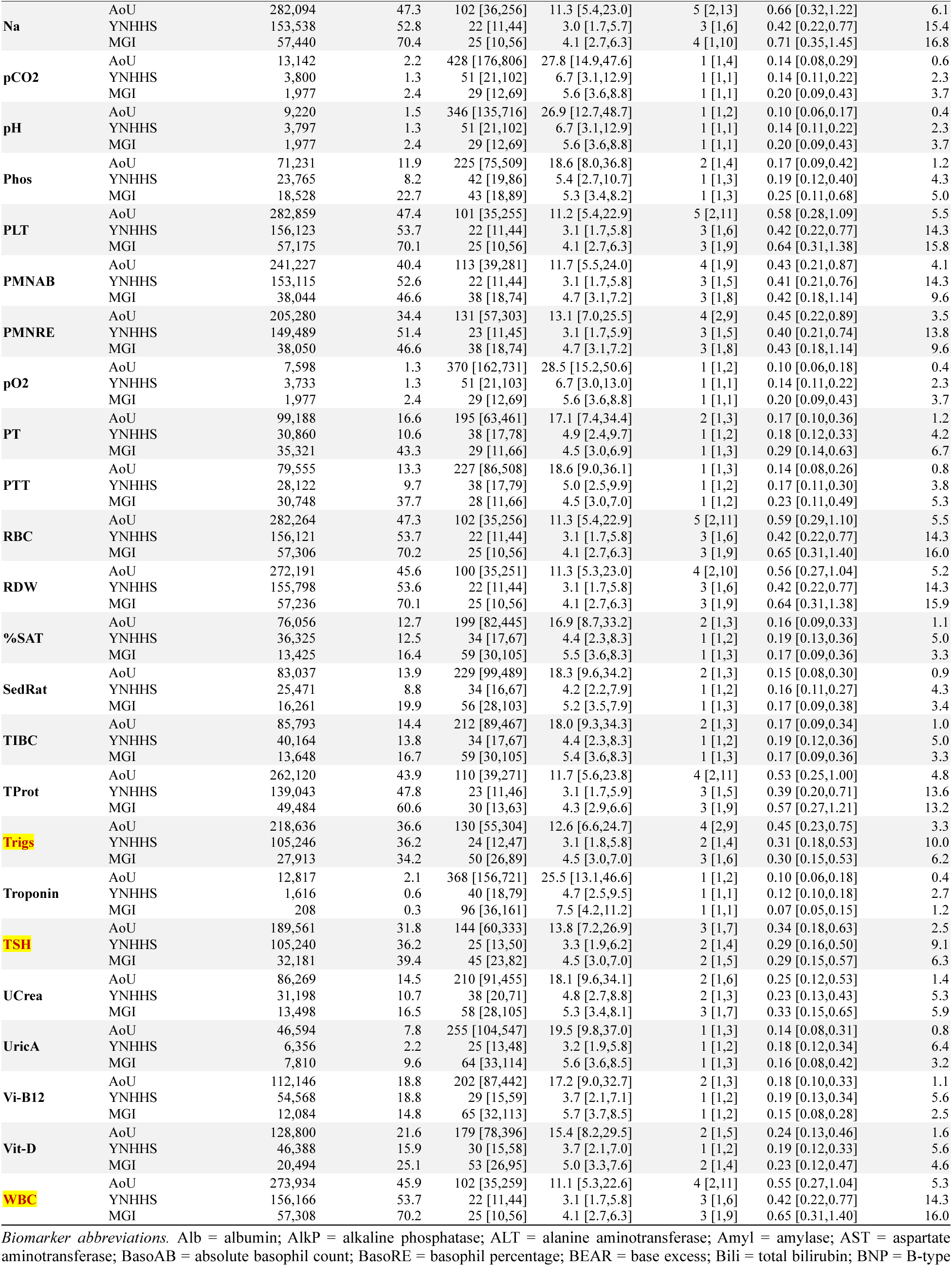
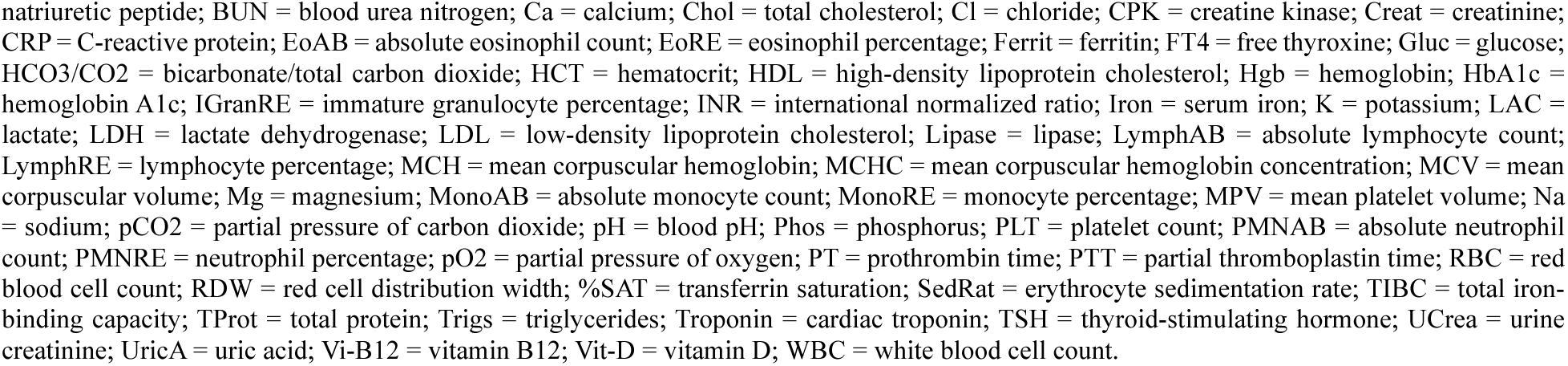
Biomarker-specific observation patterns for the 68-lab panel in AoU, YNHHS, and MGI. Patients with at least one eligible outpatient visit. Rows labelled “All” pool the entire panel among patients with eligible outpatient visits during follow-up, keeping patients with no measurements from the 68-biomarker panel as zeros; N (%) gives their number and cohort percentage, and Prop.(%) is undefined (—). For each biomarker and cohort, N is the number of patients with at least one recorded measurement of that biomarker and % is their percentage of the cohort; among these patients, Visits and Meas. are the median [Q1, Q3] eligible outpatient visits and recorded measurements per patient, Visits/Yr and Meas./Yr the corresponding median [Q1, Q3] per patient-year of follow-up, and Prop.(%) denotes the median within-patient proportion of eligible outpatient visits at which the biomarker was recorded, summarized across patients with at least one measurement.

#### Longitudinal visit characteristics stratified by covariates

Stratification shows patterns for who is seen more often (Table 4). Overall, a higher chronic disease burden was associated with a higher median number of outpatient visits per year in all three cohorts, whereas the median visit frequency by age, race, ethnicity, and neighborhood-level income differed in both magnitude and pattern across the three cohorts. The largest and most consistent differences were noted with chronic disease diagnosis: patients with diabetes had approximately twice as many outpatient visits per year as those without in AoU (12.9 vs 5.7), roughly 70% more visits in YNHHS (2.6 vs 1.5), and about 36% more visits in MGI (4.5 vs 3.3). Similar patterns were observed for cancer and for overall burden characterized by a modified Charlson comorbidity index (CCI) excluding diabetes and cancer.

**Table 4.** Outpatient-visit intensity and EHR follow-up duration by baseline covariate, per cohort. Values are median [Q1,Q3]. Visits/yr denotes outpatient visit-days per year; follow-up is measured in years.

| Variable | Level | AoU |  | YNHHS |  | MGI |  |
| --- | --- | --- | --- | --- | --- | --- | --- |
|  |  | Visits/yr | Follow-up | Visits/yr | Follow-up | Visits/yr | Follow-up |
| **Overall** | **All patients** | **6.1 [2.6, 12.3]** | **7.2 [3.1, 12.0]** | **1.7 [0.6, 3.5]** | **6.2 [3.6, 8.5]** | **3.5 [2.2, 5.6]** | **4.4 [1.6, 10.9]** |
| **Sex** | Male | 6.0 [2.4, 12.4] | 6.8 [2.8, 11.8] | 1.6 [0.6, 3.3] | 5.8 [3.1, 8.0] | 3.5 [2.1, 5.5] | 3.8 [1.4, 9.4] |
|  | Female | 6.1 [2.7, 12.2] | 7.5 [3.3, 12.2] | 1.7 [0.6, 3.7] | 6.6 [4.1, 8.9] | 3.6 [2.2, 5.6] | 5.0 [1.8, 12.2] |
| **Age** | 18-29 | 5.1 [1.8, 9.7] | 5.4 [2.3, 9.9] | 1.7 [0.7, 3.8] | 5.5 [3.0, 7.5] | 3.1 [1.9, 4.9] | 3.4 [1.5, 7.4] |
|  | 30-39 | 5.7 [2.3, 11.2] | 6.7 [2.8, 11.7] | 1.7 [0.7, 3.5] | 5.9 [3.2, 8.0] | 3.4 [2.0, 5.2] | 3.7 [1.6, 7.3] |
|  | 40-49 | 6.3 [2.9, 12.8] | 8.7 [3.8, 14.3] | 1.6 [0.6, 3.2] | 6.6 [3.8, 8.9] | 3.4 [2.1, 5.3] | 4.6 [1.7, 10.3] |
|  | 50-59 | 6.7 [3.0, 13.5] | 8.6 [3.9, 13.8] | 1.6 [0.6, 3.4] | 6.8 [4.1, 9.1] | 3.5 [2.1, 5.5] | 4.9 [1.7, 12.1] |
|  | 60-69 | 7.0 [3.0, 14.4] | 7.4 [3.3, 11.2] | 1.7 [0.6, 3.6] | 7.0 [4.4, 9.2] | 3.7 [2.3, 5.8] | 4.6 [1.6, 12.0] |
|  | 70-79 | 6.1 [2.1, 13.7] | 5.8 [2.4, 10.0] | 1.5 [0.5, 3.4] | 6.6 [3.9, 8.8] | 3.7 [2.3, 5.9] | 4.8 [1.7, 12.6] |
|  | 80+ | 5.3 [1.5, 12.6] | 3.8 [1.5, 7.9] | 1.2 [0.3, 2.9] | 5.6 [3.1, 7.5] | 3.9 [2.2, 6.4] | 4.6 [1.4, 14.0] |
| **Race** | White | 6.3 [2.9, 12.6] | 8.2 [3.8, 13.1] | 1.7 [0.7, 3.4] | 6.5 [3.8, 8.8] | 3.5 [2.2, 5.6] | 4.3 [1.6, 10.8] |
|  | Asian | 5.6 [2.6, 11.0] | 4.8 [1.9, 9.2] | 2.3 [1.1, 4.7] | 6.1 [4.1, 8.1] | 2.7 [1.6, 4.4] | 6.4 [2.7, 13.4] |
|  | Black | 6.1 [2.6, 11.7] | 6.1 [2.3, 11.2] | 1.8 [0.6, 4.0] | 6.0 [3.5, 8.1] | 3.7 [2.2, 6.1] | 5.8 [2.3, 12.6] |
|  | Other/Unknown | 5.7 [1.5, 12.2] | 5.8 [2.3, 10.4] | 1.4 [0.3, 3.3] | 5.4 [2.6, 7.6] | 3.6 [2.0, 5.8] | 3.4 [1.2, 9.0] |
| **Ethnicity** | Hispanic/Latino | 5.6 [1.5, 11.9] | 5.4 [2.1, 10.0] | 2.0 [0.7, 4.3] | 5.8 [3.2, 7.9] | 3.4 [1.8, 5.6] | 2.8 [1.2, 6.8] |
|  | Non-Hispanic | 6.1 [2.8, 12.3] | 7.6 [3.3, 12.5] | 1.7 [0.7, 3.5] | 6.4 [3.8, 8.7] | 3.6 [2.1, 5.9] | 3.1 [1.1, 8.3] |
|  | Ethnicity Unknown | 6.4 [2.5, 13.9] | 7.5 [3.2, 12.5] | 0.8 [0.0, 2.0] | 4.9 [2.0, 7.4] | 3.4 [2.2, 5.2] | 6.5 [2.9, 13.7] |
| **Neighborhood income** | Below median | 5.8 [2.5, 11.0] | 6.8 [2.8, 11.3] | 1.8 [0.7, 4.0] | 6.0 [3.5, 8.2] | 3.3 [2.0, 5.3] | 2.6 [1.0, 6.0] |
|  | At/above median | 6.7 [2.8, 13.7] | 7.7 [3.4, 12.8] | 1.6 [0.6, 3.2] | 6.3 [3.6, 8.7] | 3.7 [2.3, 5.9] | 7.4 [3.0, 15.0] |
| **BMI** | Underweight (<18.5) | 5.3 [1.9, 10.6] | 5.9 [2.4, 10.4] | 1.9 [0.8, 4.2] | 6.2 [3.9, 8.3] | 3.2 [2.1, 4.9] | 8.5 [3.2, 15.7] |
|  | Healthy (18.5-25) | 5.8 [2.6, 11.1] | 6.8 [2.9, 11.7] | 1.9 [0.9, 3.8] | 6.4 [4.1, 8.6] | 3.4 [2.1, 5.3] | 4.7 [1.8, 11.0] |
|  | Overweight (25-30) | 6.1 [2.7, 12.1] | 7.4 [3.2, 12.4] | 1.9 [0.9, 3.6] | 6.5 [4.0, 8.7] | 3.4 [2.1, 5.4] | 4.3 [1.6, 10.8] |
|  | Obese (≥30) | 6.5 [2.6, 13.5] | 7.6 [3.4, 12.1] | 2.0 [0.9, 3.9] | 6.7 [4.4, 8.9] | 3.7 [2.3, 5.9] | 4.2 [1.5, 10.5] |
| **Diabetes** | Yes | 12.9 [6.9, 23.2] | 10.8 [6.7, 16.5] | 2.6 [1.1, 5.3] | 7.6 [5.6, 9.6] | 4.5 [2.8, 7.2] | 7.0 [2.5, 15.1] |
|  | No | 5.7 [2.3, 11.0] | 6.7 [2.8, 11.3] | 1.5 [0.6, 3.2] | 6.0 [3.3, 8.3] | 3.3 [2.0, 5.2] | 3.9 [1.5, 9.5] |
| **Hypertension** | Yes | 8.3 [3.7, 16.2] | 9.9 [5.7, 14.7] | 2.2 [0.9, 4.3] | 7.3 [5.1, 9.4] | 4.0 [2.5, 6.4] | 5.5 [2.0, 13.4] |
|  | No | 4.9 [2.0, 9.0] | 4.8 [1.8, 9.4] | 1.4 [0.5, 2.9] | 5.5 [2.8, 7.8] | 3.1 [1.9, 4.9] | 3.6 [1.4, 8.5] |
| **Cancer** | Yes | 10.8 [5.5, 19.6] | 10.9 [6.9, 16.8] | 2.6 [1.1, 5.7] | 7.7 [5.5, 9.7] | 3.9 [2.4, 6.4] | 4.6 [1.7, 11.5] |
|  | No | 5.4 [2.1, 10.4] | 6.2 [2.5, 10.7] | 1.5 [0.6, 3.1] | 5.9 [3.3, 8.2] | 3.3 [2.0, 5.1] | 4.2 [1.6, 10.5] |
| **Modified CCI** | 0 | 4.8 [2.0, 8.6] | 4.9 [1.9, 9.5] | 1.4 [0.5, 2.9] | 5.5 [2.9, 7.8] | 2.8 [1.6, 4.3] | 2.8 [1.1, 6.8] |
|  | 1–2 | 7.7 [3.6, 14.4] | 9.2 [5.2, 14.0] | 2.0 [0.9, 3.9] | 7.2 [5.0, 9.3] | 3.5 [2.2, 5.2] | 5.2 [1.9, 12.4] |
|  | ≥3 | 12.2 [4.8, 23.0] | 10.8 [7.3, 16.6] | 3.5 [1.4, 7.4] | 7.7 [5.6, 9.7] | 4.6 [2.9, 7.4] | 6.2 [2.5, 13.9] |

#### Biomarker measurement characteristics

Recording frequency varied markedly by the nature and clinical use of the biomarker. Across the 68-biomarker panel in each cohort (Table 3), the proportion of patients with at least one recorded measurement (column 4) ranged from 0.1% (B-type natriuretic peptide [BNP] in YNHHS) to 71.4% (glucose in MGI). Routine chemistry and hematology biomarkers, commonly used for general assessment or longitudinal monitoring, were recorded most frequently: glucose and creatinine were recorded in roughly 48 to 71% of patients across cohorts, and blood-count biomarkers such as hemoglobin (Hgb), red blood cell count (RBC), and WBC in roughly 46 to 70%. By contrast, specialized tests for acute or high-acuity presentations were rare: lactate and blood-gas biomarkers such as pH and pO2 were recorded in fewer than 4% of patients in every cohort.

The aggregate row (labeled “All”) in Table 3 sums measurements across the 68 markers: the median number of measurements per patient was 71 in AoU, 34 in YNHHS, and 59 in MGI over follow-up, corresponding to roughly 11, 5, and 13 measurements per year, respectively. For individual biomarkers, the counts were lower: the median number of glucose measurements was 6, 3, and 4 across the three cohorts, whereas for CRP it was only 1.

At the visit level, the within-patient proportion of outpatient visits containing a biomarker (last column, Table 3) ranged from 0.4% to 19.5% across biomarker-cohort configurations. The lowest values were for amylase, troponin, pH, and pO2 in AoU (all 0.4%), whereas the highest was for glucose in MGI (19.5%), with several electrolyte and renal biomarkers in MGI measured around 17%. Although AoU patients visited far more often, each biomarker was recorded at a smaller proportion of their visits (e.g., glucose at 6.3% of visits in AoU, compared with 16.3% in YNHHS and 19.5% in MGI), reflecting their participant composition. Thus, more frequent visits did not directly translate into a higher probability of recording a given biomarker at a given visit.

### B. Which characteristics were associated with visit frequency

This analysis is related to our *second aim of modeling the visit process* and identifying its drivers.

#### Predictors of outpatient visit frequency

Table 4 presents comparisons for each characteristic separately. We then modeled outpatient visit counts using the Lin-Wei-Yang-Ying (LWYY) recurrent event model^33^, reporting unadjusted and adjusted rate ratios (RRs) by cohort in Table 5. In adjusted models, female sex was associated with higher visit rates in YNHHS and MGI but was close to null in AoU. Higher visit rates were associated with

**Table 5.** Covariate-specific outpatient visit rate ratios across AoU, YNHHS, and MGI. Estimates are visit rate ratios with 95% confidence intervals from recurrent-event models of eligible outpatient visits.

| Cohort | AoU |  | YNHHS |  | MGI |  |
| --- | --- | --- | --- | --- | --- | --- |
| Covariate | Unadjusted | Adjusted | Unadjusted | Adjusted | Unadjusted | Adjusted |
| Female (ref: male) | 0.92 (0.92, 0.93) | 1.02 (1.01, 1.03) | 0.94 (0.93, 0.96) | 1.05 (1.03, 1.06) | 1.03 (1.01, 1.05) | 1.13 (1.11, 1.14) |
| Age 40-60 (ref: <40) | 1.02 (1.01, 1.03) | 0.98 (0.97, 0.99) | 0.93 (0.92, 0.95) | 0.73 (0.72, 0.74) | 1.05 (1.03, 1.08) | 0.97 (0.95, 0.99) |
| Age ≥60 (ref: <40) | 1.24 (1.22, 1.25) | 0.99 (0.98, 1.00) | 1.07 (1.05, 1.09) | 0.62 (0.61, 0.63) | 1.20 (1.17, 1.23) | 0.94 (0.92, 0.97) |
| Black (ref: White) | 1.09 (1.08, 1.10) | 0.99 (0.98, 1.00) | 1.23 (1.21, 1.26) | 1.10 (1.06, 1.13) | 1.14 (1.10, 1.19) | 1.09 (1.06, 1.12) |
| Hispanic/Latino (ref: non-Hispanic) | 0.99 (0.96, 1.03) | 1.02 (0.99, 1.05) | 1.35 (1.32, 1.38) | 1.33 (1.30, 1.36) | 1.03 (0.92, 1.15) | 1.07 (0.99, 1.16) |
| Neighborhood-level income < median (ref: ≥median) | 0.95 (0.94, 0.96) | 0.96 (0.95, 0.96) | 1.28 (1.26, 1.29) | 1.19 (1.18, 1.21) | 0.93 (0.92, 0.95) | 0.85 (0.84, 0.86) |
| BMI≥30 (ref: <30) | 1.14 (1.13, 1.15) | 1.04 (1.03, 1.05) | 1.09 (1.07, 1.10) | 1.01 (1.00, 1.03) | 1.12 (1.10, 1.14) | 1.02 (1.01, 1.03) |
| Diabetes (ref: no diabetes) | 1.79 (1.77, 1.81) | 1.25 (1.24, 1.26) | 1.73 (1.70, 1.76) | 1.21 (1.18, 1.24) | 1.56 (1.53, 1.60) | 1.27 (1.25, 1.29) |
| Hypertension (ref: no hypertension) | 1.62 (1.60, 1.63) | 1.24 (1.23, 1.25) | 1.61 (1.59, 1.63) | 1.34 (1.32, 1.36) | 1.54 (1.52, 1.57) | 1.20 (1.18, 1.22) |
| Cancer (ref: no cancer) | 1.58 (1.57, 1.60) | 1.34 (1.33, 1.36) | 1.91 (1.88, 1.94) | 1.59 (1.55, 1.63) | 1.37 (1.35, 1.40) | 1.08 (1.06, 1.10) |
| Modified CCI 1-2 (ref: 0) | 1.27 (1.26, 1.28) | 1.42 (1.40, 1.43) | 1.26 (1.24, 1.27) | 1.42 (1.40, 1.44) | 1.03 (1.01, 1.04) | 1.38 (1.36, 1.40) |
| Modified CCI≥ 3 (ref: 0) | 1.94 (1.92, 1.96) | 1.70 (1.68, 1.72) | 2.59 (2.55, 2.64) | 2.14 (2.06, 2.21) | 1.78 (1.75, 1.81) | 1.76 (1.72, 1.80) |
| Outpatient visits in last 6 mo (vs. own avg) | 1.06 (1.06, 1.06) | 1.06 (1.05, 1.06) | 1.15 (1.14, 1.16) | 1.13 (1.12, 1.14) | 1.10 (1.08, 1.11) | 1.09 (1.08, 1.11) |

**Note.** Unadjusted estimates come from models containing one covariate at a time. Adjusted estimates compare the specified group with its reference group after adjustment for the other covariates in the model; values greater than 1 indicate more frequent outpatient visits, and values less than 1 indicate fewer outpatient visits. Because race and neighborhood-level income were not included in the same adjusted model, adjusted estimates for race, ethnicity, and all other covariates were obtained from the race model excluding neighborhood-level income, whereas the adjusted estimate for neighborhood-level income was obtained from a separate income model excluding race to avoid high correlation. Abbreviations: AoU, All of Us; CCI, Modified Charlson Comorbidity Index; MGI, Michigan Genomics Initiative; YNHHS, Yale New Haven Health System.

Hispanic or Latino ethnicity in YNHHS only, and with Black race in YNHHS and MGI; both were close to null in AoU. Obesity was associated with slightly higher visit rates in AoU but was close to null in YNHHS and MGI. Older age was associated with markedly lower adjusted visit rates in YNHHS and only slightly lower rates in AoU and MGI. Neighborhood-level income below the median was associated with higher visit rates in YNHHS but lower visit rates in AoU and MGI.

Prior diagnoses of diabetes, hypertension, and cancer remained associated with higher visit rates across all three cohorts, and comorbidity burden had the largest adjusted RR among all covariates in each cohort (Table 5). Each additional outpatient visit in the previous six months (measuring recent healthcare utilization) above the patient’s own average was also associated with a higher visit rate across all three cohorts.

### C. Which characteristics were associated with biomarker recording

This analysis illustrates *how to model the biomarker-specific observation process* and identify drivers of it.

#### Predictors of what gets measured

We focused the model-based analyses on ten biomarkers from the 68-biomarker panel (highlighted in Table 3). We fitted two models. The ever-measured model asks whether a patient has at least one recorded measurement during follow-up, a time-invariant quantity. The per-visit model asks whether that biomarker is recorded at a given outpatient visit, a time-varying quantity conditional on being seen and weighted by the estimated visit process.

#### Predictors of who gets tested at least once

We modeled the probability of at least one recorded measurement, 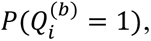 using biomarker-specific logistic regression, with unadjusted and adjusted odds ratios (ORs) in Supplementary Tables 1-3. Older age and obesity each acted consistently across cohorts and biomarkers: older age (both 40-60 and ≥60 versus <40 years) was associated with lower adjusted odds of ever being measured, except for the lipid biomarkers at age 40-60 in YNHHS, and obesity with higher odds. Associations with sex, race, ethnicity, and neighborhood-level income varied across cohorts and biomarkers. Female sex was generally associated with higher odds of having non-lipid biomarkers, particularly TSH, but showed weaker or inverse associations with lipid biomarkers. Obesity was most consistently associated with ever HbA1c measurement in all three cohorts. Below-median neighborhood income was associated with lower odds of ever measurement for every biomarker in AoU and MGI but higher odds for every biomarker in YNHHS. Hispanic or Latino ethnicity followed the same cohort pattern in AoU and YNHHS but not in MGI, and Black race was associated with lower odds for every biomarker in AoU but higher odds for every biomarker in MGI.

Clinical diagnoses showed consistent associations with ever measurement across cohorts, although patterns differed by biomarker. Diabetes showed its strongest association with ever HbA1c measurement in all three cohorts (Supplementary Tables 1-3). Hypertension was associated with higher odds of ever measurement for every biomarker in all three cohorts. Cancer showed cohort-specific associations: higher odds of ever measurement for every biomarker in AoU and YNHHS but lower odds for every biomarker in MGI.

#### Predictors of the per-visit observation model

Associations in the per-visit observation models 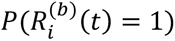 were biomarker-specific. Across all three cohorts, female sex was associated with lower adjusted odds of recording metabolic and lipid tests at a given visit, and of blood-count tests in AoU and MGI but not in YNHHS (Supplementary Figure 1). In contrast, female sex was associated with higher odds of TSH and CRP recording in all three cohorts.

Associations with other demographic and socioeconomic characteristics were less consistent across cohorts. Below-median neighborhood income showed the greatest heterogeneity in MGI, where it was associated with higher odds of recording glucose, blood-count tests, and CRP but lower odds of recording HbA1c and lipid tests; associations were weaker and more mixed in AoU and YNHHS (Figure 3). Associations with Black race were clearer in AoU and YNHHS than in MGI; in AoU and YNHHS, Black patients had higher odds of HbA1c recording but lower odds for all other labs (Supplementary Figure 1). Obesity was consistently associated with higher odds of HbA1c recording across all three cohorts, although its associations with other biomarkers were smaller and less consistent (Figure 3).

**Figure 3.**
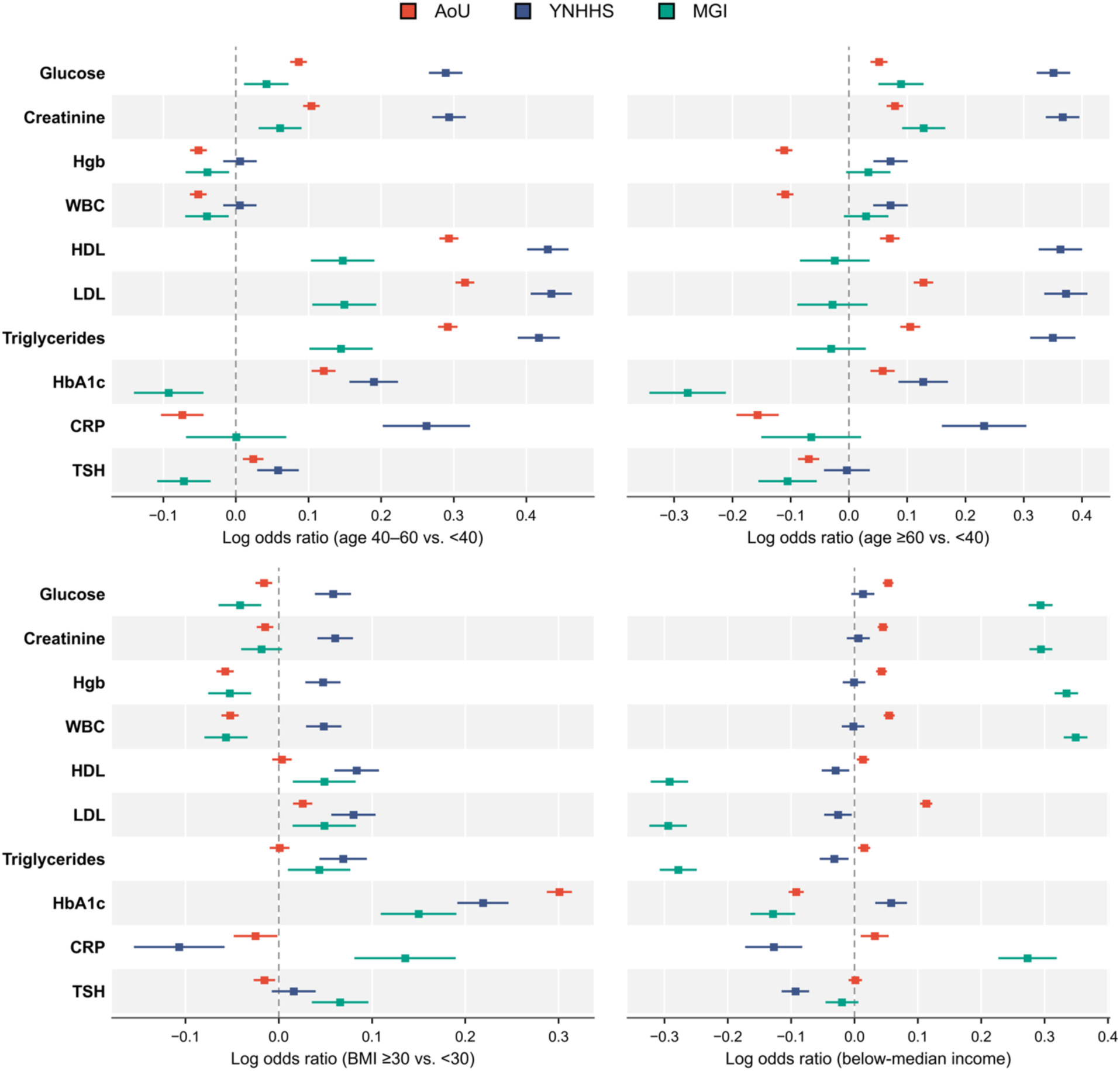
Age, body mass index (BMI) and income associations with biomarker-specific per-visit observation process. Log odds ratios with 95% confidence intervals for biomarker recording at an outpatient visit, plotted for All of Us (AoU; red), Yale New Haven Health System (YNHHS; blue) and the Michigan Genomics Initiative (MGI; green). Estimates correspond to age categorized into three groups (top left and right), BMI≥30 versus BMI <30 (bottom left), and below-median versus at-or-above-median annual neighborhood-level household income (bottom right), conditional on the other covariates in the biomarker-specific generalized estimating equation model. See Methods for model specification.

By contrast, associations with prior diagnoses and recent measurement history were more consistent across cohorts. Diabetes was more strongly associated with HbA1c recording than with glucose recording (Figure 4). Hypertension was associated with higher odds of recording glucose, creatinine, HbA1c, and lipid tests, whereas cancer was associated with higher odds of recording chemistry and blood-count tests but lower odds of recording HbA1c, lipids, and CRP. Direction and magnitude of associations with recent measurement history were also biomarker-specific: prior CRP measurement was strongly associated with repeat CRP recording, whereas prior lipid measurement was associated with lower odds of repeat recording in AoU but not in YNHHS or MGI (Supplementary Figure 2). Unadjusted per-visit ORs are provided in Supplementary Tables 4-6.

**Figure 4.**
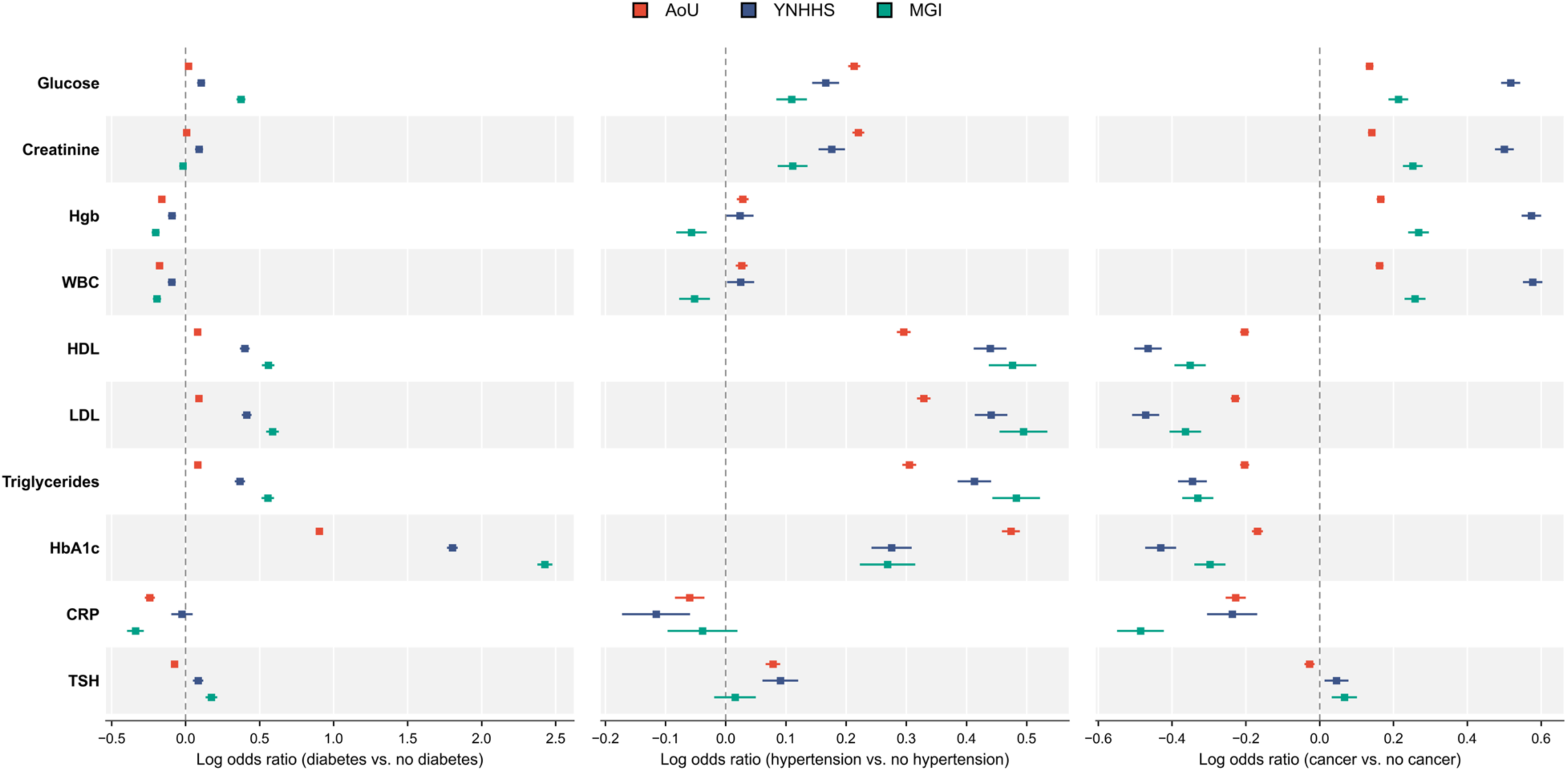
Comorbidity-related associations with biomarker-specific observation. Log odds ratios with 95% confidence intervals for biomarker recording at an outpatient visit, plotted for All of Us (AoU; red), Yale New Haven Health System (YNHHS; blue) and the Michigan Genomics Initiative (MGI; green). Estimates correspond to diabetes (left), hypertension (center), and cancer status (right), conditional on the other covariates in the biomarker-specific generalized estimating equation model. See Methods for model specification.

## Discussion

### Main Finding

Across three EHR-linked cohorts, we characterized two distinct recording processes underlying a data stream of longitudinal biomarker values: the occurrence of a recorded outpatient visit and biomarker recording conditional on that visit. Prior methods for irregular longitudinal data have largely focused on the visit process^19–23^, whereas our results show that the factors associated with biomarker recording at a visit may differ from those associated with visit frequency.

#### What predicts visits?

Across cohorts, diabetes, hypertension, cancer, comorbidity burden, and recent outpatient utilization were the most consistent predictors of higher visit rates, whereas demographic and socioeconomic associations varied. Disease burden and utilization history are therefore reasonable core predictors for visit-process models, whereas other terms require evaluation in the specific study population.

#### What predicts lab tests?

Conditional on a recorded outpatient visit, the probability of biomarker recording varied by biomarker, clinical history, and recent measurement history. Female sex was associated with lower recording of metabolic, lipid, and blood-count biomarkers but higher recording of TSH and CRP; diabetes was more strongly associated with HbA1c than glucose; and cancer with more chemistry and blood-count recording but less lipid and HbA1c recording. Recent CRP and lipid measurements in AoU were associated with subsequent recording in opposite directions. These patterns are compatible with differences in clinical indication and monitoring schedules; glycemic and lipid guidelines, for example, recommend different intervals between repeat tests^30,31^. Observation models should therefore be fitted separately for each biomarker.

### Contrasting the ever-measured and per-visit models

The two models sometimes gave associations in opposite directions for the same predictor and biomarker. In AoU, for example, modified CCI ≥3 versus 0 was associated with higher odds that HDL, LDL, and triglycerides were ever measured (Supplementary Tables 1-3). In contrast, the same comparison was associated with lower odds that these biomarkers were recorded at a given outpatient visit, with similar opposing patterns observed in YNHHS and MGI.

Ever-measured and per-visit analyses target different probabilities, so their associations need not have the same direction. The negative per-visit association does not imply fewer lipid measurements over follow-up; rather, it indicates lower conditional odds that a given outpatient visit contained a lipid measurement. One plausible explanation is that patients with greater comorbidity burden accumulated outpatient visits for other needs, whereas lipid monitoring remained intermittent^30^. Total visits may therefore increase more than lipid-testing visits, lowering per-visit recording while increasing opportunities for at least one measurement. A model for what gets measured should account for visit frequency and time since the biomarker was last measured. Because associations differed by cohort, the model should be refitted in each setting rather than borrowed from another.

#### What is the consequence for downstream association tasks?

Because the visit and observation processes are driven by different factors, removing one but not the other does not guarantee an unbiased association when the biomarker values are treated as a longitudinal outcome and regressed against genetic or environmental risk factors. Adjusting for the visit process^19–23^ alone can make matters worse. For the sake of completeness, we conducted a simulation study mimicking a subset of our previous work^25^ (summarized in Supplementary Section 3 with results in Supplementary Figures 3 and 4). We compared ten methods (five naïve, three IP-aware and two adjusting for both IP and IO) for longitudinal association analysis. Naïve analyses that ignore IP and IO (a linear mixed model, or a linear regression on patient’s mean, median or summary biomarker value) were all biased for the association coefficient, by about 12% for the linear mixed model. Adjusting for IP but not IO failed to remove this bias and increased the percentage bias to about 28%. Only analyses that account for both IP and IO recovered the target association and produced nearly unbiased estimates. The visit and observation models characterized here are therefore not merely descriptive: they supply the weights and propensities a downstream analysis needs, and methods that use both stages already exist with usable software and should be used in a longitudinal association analysis^25^.

### Limitations

*First*, the visit and observation models were estimated separately for ease of implementation, although shared unmeasured health factors may drive both processes and the biomarker; joint modeling is therefore more principled^25,26^. Biomarkers always co-measured on a panel could also be modeled jointly. *Second*, we assumed non-informative termination and did not model death; if death or loss to EHR follow-up depends on health status that also drives visiting and observation, methods for informative censoring or terminal events are required^36–40^. *Third*, the models condition on recorded covariates and cannot capture dependence on the unobserved current biomarker value, unrecorded symptoms, clinical suspicion, or latent health states, so residual bias may remain^41–43^. *Fourth*, we observed outpatient visits and biomarker results but not test orders or completion and therefore could not distinguish tests not ordered from those ordered but not completed or captured. *Finally*, predictors and functional forms were prespecified for interpretability and cross-cohort comparability. Omitted predictors (such as medication history), nonlinearities, or interactions could misspecify either model; penalized regression, machine learning, or EHR foundation-model^44^ embeddings may better capture these features. Thus, the fitted models describe associations of observed covariates with each process rather than the complete recording mechanism.

Single-system EHRs provide an incomplete snapshot of a patient’s health, omitting physiology between visits and care received elsewhere. Our restriction to laboratory measurements attributable to outpatient visits may also omit biomarker monitoring occurring in other care settings. Cross-system aggregation, participant-mediated EHR linkage, and consumer-device data^45^ could provide a more comprehensive view of patients’ health trajectories.

Our framework and empirical exposition suggest that association analysis with longitudinal EHR biomarkers as outcomes should characterize both visit and observation processes. The measured and unmeasured drivers of these processes vary across cohorts and biomarkers and should therefore be evaluated within each study setting.

## Methods

### Study and data access approval

All research in this study was conducted under the applicable institutional review board approvals and data-use agreements. The All of Us Research Program protocol was reviewed by the All of Us Institutional Review Board, and patients provided informed consent, including authorization for the research use of electronic health record data. All of Us data were accessed by credentialed researchers through approved Researcher Workbench workspaces under Yale University’s institutional All of Us Data Use and Registration Agreement. Use of Yale New Haven Health System data was approved by the Yale University Institutional Review Board under protocol 2000040898. Michigan Genomics Initiative data were used under University of Michigan Institutional Review Board record HUM00177982 and the Yale–Michigan Federal Demonstration Partnership Data Transfer and Use Agreement 26-UFA02513. All analyses were conducted in accordance with the applicable institutional approvals, patient privacy protections, controlled-access policies and data-use agreements.

### Study design and data sources

All of Us (AoU). The All of Us Research Program is a National Institutes of Health initiative launched in 2018 with the goal of enrolling more than one million US adults, recruited through a combination of open, direct-to-patient invitations and a national network of healthcare provider organizations^27^. Engagement has deliberately oversampled groups historically underrepresented in biomedical research, targeting diversity across age, sex, race and ethnicity, gender identity, sexual orientation, disability status, healthcare access, income, educational attainment, and geographic location^27^, with patient-reported information linked to electronic health records contributed from many care settings nationally. The analytic subset comprised n = 599,423 patients with sociodemographic data and linked ICD-9-CM/ICD-10-CM diagnoses and lab measurements, drawn from the Controlled Tier of All of Us Curated Data Repository version 9 (CDRv9; C2025Q4R6); analyzed records spanned 2000 through the January 1, 2025, data cutoff.

Yale New Haven Health System (YNHHS). YNHHS is the largest health system in Connecticut, providing inpatient and outpatient care through several member hospitals and an extensive ambulatory network across Connecticut and the surrounding region, all on a unified Epic electronic health record. Person-level clinical data are assembled for research through an integrated data-science platform that consolidates the system’s EHR, lab, and administrative data into a research-ready repository^28^. Because participation follows from receipt of care rather than from consent into a genomics study, the cohort reflects the multi-ancestry population served by the health system in southern New England rather than a sample shaped by a specific recruitment protocol. The analytic subset comprised n = 319,666 patients with sociodemographic data and linked ICD diagnoses and lab measurements; records spanned 2012 to 2025.

Michigan Genomics Initiative (MGI). MGI is a single-institution biobank at Michigan Medicine (University of Michigan) that began enrolling adults in 2012, primarily at preoperative encounters for procedures requiring anesthesia^29^. It subsequently expanded to additional specialty sub-cohorts, including metabolism, endocrinology and diabetes (MEND) and mental health (MHB) clinic populations and a wearables cohort enriched for hypertension (MIPACT)^29^. Because recruitment is concentrated in surgical and specialty settings, MGI comprises an older and more clinically selected population, carrying a higher burden of chronic disease than the general population. The analytic subset comprised n = 82,372 patients with sociodemographic data and linked ICD-9-CM/ICD-10-CM diagnoses and lab measurements, drawn from an earlier data extract (records through 2019) in which event times were recorded as days since birth rather than calendar dates.

### Cohort, follow-up and biomarker linkage

Within each cohort, the analysis population comprised patients aged 18 years or older at cohort entry with at least one clinical event in the linked EHR, where a clinical event was any dated encounter of any type, laboratory measurement, or procedure record. Age was defined at cohort entry as the difference between the calendar year of the patient’s first observed clinical event and their birth year. Details of cohort construction and measurement attribution are shown in Supplementary Figures 5 and 6, respectively.

Follow-up was defined using standard Observational Health Data Sciences and Informatics (OHDSI) conventions for EHR data: an observation period ended after an extended gap in EHR activity and a new one began when the patient returned, so that time during which patients were no longer receiving care within the system was not counted as follow-up (548-day gap threshold; each period extended 60 days beyond its last recorded event)^46^. Follow-up started 90 days after each patient’s first recorded clinical event; this initial window did not contribute follow-up time, and visits within it were not counted. Outpatient visits were identified using OMOP standard outpatient concepts (descendants of concept_id 9202), excluding emergency department and inpatient visits, with multiple records on the same calendar day collapsed into a single visit day. Biomarkers were identified using harmonized LOINC codes; values were converted to standard reference units and implausible values outside biomarker-specific clinical bounds were set to missing. The outcome for the conditional observation-process analysis was a binary indicator for whether each biomarker was linked to a given outpatient visit. Time-varying covariates entering the visit and observation models were updated at each visit using a strictly-before convention for diagnosis.

### Visit-process model

We modeled outpatient visits as a recurrent-event process using the Lin-Wei-Yang-Ying (LWYY) marginal proportional-rate model^33^. For patient *i*, let *N_i_*(*t*) denote the cumulative number of eligible outpatient visit days by time *t* after baseline. Multiple outpatient records on the same calendar day were counted as a single visit day. We assumed the multiplicative model,

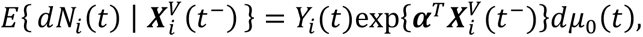

where 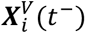 is the covariate vector just before time *t*, so that every covariate uses only information available strictly before *t*; *Y_i_*(*t*) is the at-risk indicator, equal to one when patient *i* is under eligible follow-up at time *t* and 0 otherwise, so that visits are counted only within eligible observation periods; ***α*** indexes associations between the covariates and outpatient visits; and *dμ*_0_(*t*) is an unspecified baseline mean-rate function. Exponentiated coefficients were interpreted as visit rate ratios, with values greater than 1 indicating more frequent outpatient visit among patients with that characteristic, conditional on the other covariates. Regression coefficients were estimated separately within each cohort using the survival package^47^ (version 3.8-9) in R (version 4.5.1), with Breslow’s method for tied event times, and robust sandwich standard errors clustered by patient. Models were fitted on complete cases.

For the cross-cohort analysis, 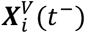 included age as a three-level categorical variable (<40 [reference], 40-60, ≥60), sex, race, ethnicity, an indicator for obesity (recorded body mass index [BMI]≥30 kg/m^2^), an indicator for below-median neighborhood-level annual household income, defined as the median household income of the patient’s residential three-digit ZIP code area and dichotomized at the median of that variable, and time-varying indicators for diabetes, hypertension and cancer, defined from SNOMED concept sets and switching on at the first recorded diagnosis. The modified Charlson Comorbidity Index was a weighted sum of the standard Charlson components, excluding diabetes and cancer, updated as each component was first recorded, and was included as a categorical variable with levels 0, 1-2, and ≥3, with 0 as the reference level. We also included a time-varying summary of recent outpatient use. At each time *t*, this variable was defined as the number of eligible visit days in the preceding six months, excluding time *t*, minus the patient’s own running average of the same six-month count over earlier follow-up.

### Ever-measured model

In addition to the per-visit model, we fitted an ever-measured model for each biomarker. For patient *i* and biomarker *b*, 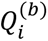 was defined as 1 if patient *i* had at least one recorded measurement of biomarker *b* over follow-up and 0 otherwise. We modeled 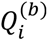 using logistic regression with the same person-level covariates. Because this outcome is time-invariant, the prior-measurement term was not included.

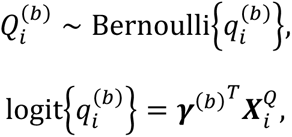

where 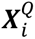 is the covariate vector, and ***γ***^(*b*)^ is the vector of biomarker-specific regression coefficients. For the cross-cohort analysis, 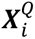 included categorized sex, age, race, ethnicity, obesity, below-median neighborhood-level annual household income, indicators for diabetes, hypertension, and cancer, and the modified CCI. Exponentiated coefficients were interpreted as ORs for having at least one recorded measurement of the biomarker during the observed follow-up period, conditional on the other covariates. These ORs therefore describe cumulative recording during the available follow-up and may reflect both measurement practices and the duration of EHR observation. A separate model was fitted for each biomarker within each cohort.

### Per-visit observation-process model

Conditional on an outpatient visit, we modeled whether each biomarker was recorded at that visit, weighting by the visit process so that the estimates describe recording for the population that could potentially visit rather than only the subset observed to visit. For biomarker *b*, patient *i*, and outpatient visits occurring at time *t*, let 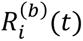 equal 1 if biomarker *b* was recorded at that visit and 0 otherwise. We used a biomarker-specific generalized estimating equation (GEE)^34^ with binomial family, logit link, and independent working correlation:

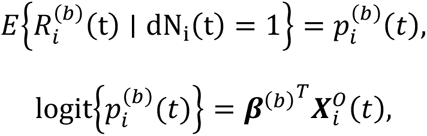

where 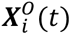 is the covariate vector evaluated at visit time *t*, and ***β***^(*b*)^ is the vector of biomarker-specific regression coefficients. For the cross-cohort analysis, 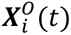 included categorized sex, age, race, ethnicity, obesity, below-median neighborhood-level annual household income, time-varying indicators for diabetes, hypertension and cancer, the modified CCI, the time-varying summary of recent outpatient use defined above, and the number of prior recorded measurements of biomarker *b* during the six months before the visit.

The model was fitted using weights derived from the visit-process model to account for selective visit occurrence. An independent working correlation was used, and within-patient correlation across repeated visits was accommodated through the empirical sandwich variance estimator clustered by patient. Exponentiated coefficients were interpreted as ORs for recording biomarker *b* at a given outpatient visit, conditional on the other covariates and after accounting for the visit process. A separate model was fitted for each biomarker within each cohort. The per-visit GEE and the ever-measured logistic model target two distinct probabilities: the probability that a biomarker is recorded at a given outpatient visit and the probability that a patient has at least one recorded measurement of that biomarker during the observed follow-up period.

## Data availability

All of Us data are available to authorized researchers through the Researcher Workbench, subject to the program’s standard access requirements, including coverage under an institutional Data Use and Registration Agreement and completion of the required registration and training. Analyses used the Controlled Tier Curated Data Repository version 9 (C2025Q4R6). Individual-level YNHHS and MGI data cannot be publicly shared or redistributed by the authors because they are governed by institutional approvals and data-use agreements. Source data are provided with this paper.

## References

1. Bycroft, C. et al. The UK Biobank resource with deep phenotyping and genomic data. Nature 562, 203–209 (2018).

2. Bick, A. G. et al. Genomic data in the All of Us Research Program. Nature 627, 340–346 (2024).

3. Diogo, D. et al. Phenome-wide association studies across large population cohorts support drug target validation. Nat. Commun. 9, 4285 (2018).

4. Goldstein, J. A. et al. LabWAS: Novel findings and study design recommendations from a meta-analysis of clinical labs in two independent biobanks. PLOS Genet. 16, e1009077 (2020).

5. McGee, G., Haneuse, S., Coull, B. A., Weisskopf, M. G. & Rotem, R. S. On the Nature of Informative Presence Bias in Analyses of Electronic Health Records. Epidemiology 33, 105–113 (2022).

6. Harton, J., Mitra, N. & Hubbard, R. A. Informative presence bias in analyses of electronic health records-derived data: a cautionary note. J. Am. Med. Inform. Assoc. JAMIA 29, 1191–1199 (2022).

7. Sisk, R. et al. Informative presence and observation in routine health data: A review of methodology for clinical risk prediction. J. Am. Med. Inform. Assoc. JAMIA 28, 155–166 (2021).

8. Goldstein, B. A., Phelan, M., Pagidipati, N. J. & Peskoe, S. B. How and when informative visit processes can bias inference when using electronic health records data for clinical research. J. Am. Med. Inform. Assoc. JAMIA 26, 1609–1617 (2019).

9. Carrero, J. J. et al. Defining measures of kidney function in observational studies using routine health care data: methodological and reporting considerations. Kidney Int. 103, 53–69 (2023).

10. Goldstein, B. A., Bhavsar, N. A., Phelan, M. & Pencina, M. J. Controlling for Informed Presence Bias Due to the Number of Health Encounters in an Electronic Health Record. Am. J. Epidemiol. 184, 847–855 (2016).

11. Fiori, K. P. et al. Unmet Social Needs and No-Show Visits in Primary Care in a US Northeastern Urban Health System, 2018–2019. Am. J. Public Health 110, S242–S250 (2020).

12. Ashman, J., Santo, L. & Okeyode, T. Characteristics of Office-Based Physician Visits, 2018. https://stacks.cdc.gov/view/cdc/105509 (2021) doi:10.15620/cdc:105509.

13. Wang, J. et al. Detection Bias in EHR-Based Research on Clinical Exposures and Dementia. *JAMA Netw*. Open 8, e256637 (2025).

14. Colwell, R. L., Narayan, A. K. & Ross, A. B. Patient Race or Ethnicity and the Use of Diagnostic Imaging: A Systematic Review. J. Am. Coll. Radiol. JACR 19, 521–528 (2022).

15. Silva, E. L. et al. Polycystic Ovary Syndrome Underdiagnosis Patterns by Individual-level and Spatial Social Vulnerability Measures. J. Clin. Endocrinol. Metab. 110, 1657–1666 (2025).

16. Chavez-Yenter, D. et al. Association of Disparities in Family History and Family Cancer History in the Electronic Health Record With Sex, Race, Hispanic or Latino Ethnicity, and Language Preference in 2 Large US Health Care Systems. JAMA Netw. Open 5, e2234574 (2022).

17. Xu, X., Chen, L., Nunez-Smith, M., Clark, M. & Wright, J. D. Racial disparities in diagnostic evaluation of uterine cancer among Medicaid beneficiaries. J. Natl. Cancer Inst. 115, 636–643 (2023).

18. Schaffer, J. M. et al. Insurance-related differences in Chronic Conditions Data Warehouse comorbidities of Medicare beneficiaries. Am. J. Manag. Care 31, e336–e346 (2025).

19. Lin, H., Scharfstein, D. O. & Rosenheck, R. A. Analysis of Longitudinal Data with Irregular, Outcome-Dependent Follow-Up. J. R. Stat. Soc. Ser. B Stat. Methodol. 66, 791–813 (2004).

20. Buzkova, P. & Lumley, T. Longitudinal data analysis for generalized linear models with follow-up dependent on outcome-related variables. Can. J. Stat. 35, 485–500 (2007).

21. Liang, Y., Lu, W. & Ying, Z. Joint modeling and analysis of longitudinal data with informative observation times. Biometrics 65, 377–384 (2009).

22. Chen, Y., Ning, J. & Cai, C. Regression analysis of longitudinal data with irregular and informative observation times. Biostatistics 16, 727–739 (2015).

23. Pullenayegum, E. M. & Lim, L. S. Longitudinal data subject to irregular observation: A review of methods with a focus on visit processes, assumptions, and study design. Stat. Methods Med. Res. 25, 2992–3014 (2016).

24. Anthopolos, R., Wei, Y. & Chen, Q. Modeling Heterogeneity and Missing Data of Multiple Longitudinal Outcomes in Electronic Health Records. Preprint at 10.48550/arXiv.2103.11170 (2021).

25. Yang, C.-H., Shi, X. & Mukherjee, B. Joint Modeling of Longitudinal EHR Data with Shared Random Effects for Informative Visiting and Observation Processes. Preprint at 10.48550/arXiv.2602.15374 (2026).

26. Du, J., Shi, X. & Mukherjee, B. A new statistical approach for joint modeling of longitudinal outcomes measured in electronic health records with clinically informative presence and observation processes. Preprint at 10.48550/arXiv.2410.13113 (2025).

27. The All of Us Research Program Investigators. The “All of Us” Research Program. N. Engl. J. Med. 381, 668–676 (2019).

28. McPadden, J. et al. Health Care and Precision Medicine Research: Analysis of a Scalable Data Science Platform. J. Med. Internet Res. 21, e13043 (2019).

29. Zawistowski, M. et al. The Michigan Genomics Initiative: A biobank linking genotypes and electronic clinical records in Michigan Medicine patients. Cell Genomics 3, (2023).

30. Grundy, S. M. et al. 2018 AHA/ACC/AACVPR/AAPA/ABC/ACPM/ADA/AGS/APhA/ASPC/NLA/PCNA Guideline on the Management of Blood Cholesterol: A Report of the American College of Cardiology/American Heart Association Task Force on Clinical Practice Guidelines. Circulation 139, e1082–e1143 (2019).

31. American Diabetes Association Professional Practice Committee. 6. Glycemic Goals and Hypoglycemia: Standards of Care in Diabetes-2025. Diabetes Care 48, S128–S145 (2025).

32. Gulhar, R., Ashraf, M. A. & Jialal, I. Physiology, Acute Phase Reactants. in StatPearls (StatPearls Publishing, Treasure Island (FL), 2026).

33. Lin, D. Y., Wei, L. J., Yang, I. & Ying, Z. Semiparametric regression for the mean and rate functions of recurrent events. J. R. Stat. Soc. Ser. B Stat. Methodol. 62, 711–730 (2000).

34. Liang, K.-Y. & Zeger, S. L. Longitudinal Data Analysis Using Generalized Linear Models. Biometrika 73, 13–22 (1986).

35. Dennis, J. K. et al. Clinical laboratory test-wide association scan of polygenic scores identifies biomarkers of complex disease. Genome Med. 13, 6 (2021).

36. Wang, M.-C., Qin, J. & Chiang, C.-T. Analyzing Recurrent Event Data With Informative Censoring. J. Am. Stat. Assoc. 96, 10.1198/016214501753209031 (2001).

37. Robins, J. M., Rotnitzky, A. & Zhao, L. P. Analysis of Semiparametric Regression Models for Repeated Outcomes in the Presence of Missing Data. J. Am. Stat. Assoc. 90, 106–121 (1995).

38. Ghosh, D. & Lin, D. Y. Marginal Regression Models for Recurrent and Terminal Events. Stat. Sin. 12, 663–688 (2002).

39. Liu, L., Wolfe, R. A. & Huang, X. Shared frailty models for recurrent events and a terminal event. Biometrics 60, 747–756 (2004).

40. Liu, L., Huang, X. & O’Quigley, J. Analysis of Longitudinal Data in the Presence of Informative Observational Times and a Dependent Terminal Event, with Application to Medical Cost Data. Biometrics 64, 950–958 (2008).

41. Rubin, D. B. Inference and Missing Data. Biometrika 63, 581–592 (1976).

42. Little, R. J. A. & Rubin, D. B. Statistical Analysis with Missing Data. (John Wiley & Sons, 2019).

43. Mealli, F. & Rubin, D. B. Clarifying missing at random and related definitions, and implications when coupled with exchangeability. Biometrika 102, 995–1000 (2015).

44. Wornow, M. et al. The shaky foundations of large language models and foundation models for electronic health records. Npj Digit. Med. 6, 135 (2023).

45. Pedroso, A. F. & Khera, R. Leveraging AI-enhanced digital health with consumer devices for scalable cardiovascular screening, prediction, and monitoring. *Npj Cardiovasc*. Health 2, 34 (2025).

46. Hripcsak, G. et al. Observational Health Data Sciences and Informatics (OHDSI): Opportunities for Observational Researchers. Stud. Health Technol. Inform. 216, 574–578 (2015).

47. Therneau, T. M. survival: Survival Analysis. 3.8-9 10.32614/CRAN.package.survival (2001).

