## Supplementary Material for "Who seeks care, and what gets measured? Understanding the distinct mechanisms behind visit and observation processes in multi-center electronic health records"

### Contents

**S1. Ever-measured models**

**Supplementary Table 1.** All of Us: ever-measured logistic model.
**Supplementary Table 2.** Yale New Haven Health System: ever-measured logistic model.
**Supplementary Table 3.** Michigan Genomics Initiative: ever-measured logistic model.

**S2. Per-visit observation models**

**Supplementary Table 4.** All of Us: univariate per-visit observation model.
**Supplementary Table 5.** Yale New Haven Health System: univariate per-visit observation model.
**Supplementary Table 6.** Michigan Genomics Initiative: univariate per-visit observation model.
**Supplementary Figure 1.** Sex, race and ethnicity associations with biomarker-specific observation.
**Supplementary Figure 2.** Comorbidity burden and prior-measurement associations with biomarker-specific observation.

**S3. Simulation study**

**Simulation design.**
**Estimators evaluated.**
**Simulation results.**
**Supplementary Figure 3.** Bias of estimators in the simulation study.
**Supplementary Figure 4.** Root mean squared error (RMSE) of estimators in the simulation study.

**S4. Cohort definition and measurement attribution**

**Supplementary Figure 5.** Definition of the analytic cohort.
**Supplementary Figure 6.** Mapping of laboratory measurements to outpatient visits.

### S1. Ever-measured models

This section presents participant-level logistic models for whether each biomarker was recorded at least once during follow-up. Supplementary Tables 1-3 report cohort-specific univariate and mutually adjusted odds ratios for All of Us, Yale New Haven Health System and the Michigan Genomics Initiative, respectively.

**Supplementary Table 1 | All of Us: ever-measured logistic model.** Odds ratios (95% CI) by covariate, univariate (unadjusted) and adjusted. Because race/ethnicity and neighborhood income are collinear, the adjusted estimates come from two models: the race model (Black and Hispanic indicators; income excluded) provides the adjusted estimates for race, ethnicity and all other covariates, whereas a separate income model (below-median income indicator; race/ethnicity excluded) provides the adjusted estimate for income. CCI, modified Charlson Comorbidity Index (excludes diabetes and cancer). Reference groups: male, age <40, White, non-Hispanic, at/above-median income, BMI <30, absence of each condition.

| Biomarker | Female | Age 40-60 | Age ≥60 | Black | Hispanic | Low income | Obese | Diabetes | Hypertension | Cancer | CCI 1-2 | CCI ≥3 |
|---|---|---|---|---|---|---|---|---|---|---|---|---|
| **Univariate (unadjusted)** | | | | | | | | | | | | |
| **Glucose** | 1.03 (1.02, 1.05) | 1.55 (1.53, 1.57) | 0.67 (0.66, 0.68) | 0.52 (0.52, 0.53) | 0.67 (0.64, 0.70) | 0.68 (0.68, 0.69) | 1.78 (1.76, 1.80) | 7.96 (7.70, 8.23) | 8.78 (8.65, 8.91) | 8.64 (8.45, 8.84) | 4.75 (4.67, 4.84) | 9.30 (9.05, 9.56) |
| **Creatinine** | 1.03 (1.02, 1.04) | 1.57 (1.55, 1.59) | 0.68 (0.67, 0.69) | 0.53 (0.52, 0.53) | 0.67 (0.64, 0.69) | 0.67 (0.66, 0.67) | 1.79 (1.77, 1.82) | 9.07 (8.76, 9.40) | 9.50 (9.36, 9.65) | 9.00 (8.79, 9.21) | 5.02 (4.93, 5.11) | 9.68 (9.41, 9.96) |
| **Hgb** | 1.13 (1.11, 1.14) | 1.45 (1.44, 1.47) | 0.63 (0.63, 0.64) | 0.53 (0.52, 0.53) | 0.70 (0.68, 0.73) | 0.72 (0.71, 0.73) | 1.76 (1.74, 1.78) | 6.33 (6.14, 6.53) | 7.67 (7.56, 7.78) | 7.55 (7.39, 7.71) | 4.53 (4.46, 4.61) | 8.51 (8.29, 8.74) |
| **WBC** | 1.14 (1.13, 1.15) | 1.45 (1.44, 1.47) | 0.64 (0.63, 0.65) | 0.55 (0.54, 0.56) | 0.70 (0.67, 0.73) | 0.74 (0.73, 0.75) | 1.78 (1.75, 1.80) | 6.48 (6.29, 6.68) | 7.72 (7.61, 7.84) | 7.64 (7.48, 7.81) | 4.44 (4.37, 4.51) | 8.53 (8.31, 8.76) |
| **HDL** | 0.97 (0.95, 0.98) | 1.67 (1.65, 1.69) | 0.62 (0.62, 0.63) | 0.56 (0.55, 0.57) | 0.59 (0.56, 0.61) | 0.66 (0.66, 0.67) | 1.61 (1.59, 1.63) | 6.17 (6.00, 6.33) | 5.65 (5.57, 5.73) | 4.47 (4.40, 4.55) | 3.16 (3.11, 3.21) | 4.00 (3.92, 4.08) |
| **LDL** | 0.99 (0.98, 1.00) | 1.68 (1.66, 1.70) | 0.63 (0.62, 0.64) | 0.58 (0.57, 0.59) | 0.56 (0.54, 0.59) | 0.77 (0.77, 0.78) | 1.63 (1.61, 1.65) | 7.04 (6.84, 7.23) | 5.90 (5.82, 5.98) | 4.35 (4.27, 4.43) | 3.24 (3.19, 3.29) | 3.95 (3.88, 4.03) |
| **Triglycerides** | 0.96 (0.95, 0.97) | 1.67 (1.65, 1.69) | 0.64 (0.63, 0.65) | 0.56 (0.55, 0.57) | 0.59 (0.57, 0.62) | 0.66 (0.65, 0.67) | 1.60 (1.58, 1.62) | 6.24 (6.08, 6.41) | 5.77 (5.70, 5.86) | 4.51 (4.43, 4.59) | 3.18 (3.13, 3.23) | 4.07 (3.99, 4.16) |
| **HbA1c** | 0.90 (0.88, 0.91) | 1.62 (1.59, 1.64) | 0.69 (0.68, 0.70) | 0.78 (0.77, 0.79) | 0.70 (0.67, 0.73) | 0.62 (0.62, 0.63) | 2.03 (2.00, 2.06) | 10.29 (10.02, 10.56) | 6.19 (6.10, 6.28) | 3.46 (3.40, 3.52) | 2.66 (2.62, 2.70) | 4.33 (4.24, 4.41) |
| **CRP** | 1.22 (1.19, 1.24) | 1.50 (1.47, 1.52) | 0.68 (0.67, 0.70) | 0.61 (0.60, 0.63) | 0.69 (0.65, 0.74) | 0.78 (0.77, 0.79) | 1.70 (1.67, 1.74) | 3.17 (3.09, 3.25) | 4.09 (4.01, 4.16) | 3.13 (3.06, 3.19) | 2.57 (2.52, 2.61) | 4.36 (4.27, 4.45) |
| **TSH** | 1.25 (1.23, 1.27) | 1.47 (1.45, 1.49) | 0.65 (0.64, 0.66) | 0.53 (0.52, 0.54) | 0.72 (0.69, 0.75) | 0.69 (0.69, 0.70) | 1.53 (1.51, 1.55) | 3.83 (3.74, 3.92) | 4.41 (4.35, 4.48) | 4.11 (4.04, 4.18) | 2.90 (2.86, 2.95) | 3.47 (3.41, 3.54) |
| **Adjusted (mutually adjusted)** | | | | | | | | | | | | |
| **Glucose** | 1.16 (1.15, 1.18) | 0.75 (0.74, 0.76) | 0.30 (0.29, 0.30) | 0.34 (0.33, 0.35) | 0.69 (0.66, 0.72) | 0.66 (0.65, 0.67) | 1.14 (1.12, 1.16) | 2.10 (2.01, 2.19) | 4.93 (4.83, 5.02) | 5.15 (5.01, 5.30) | 3.92 (3.84, 4.00) | 5.63 (5.44, 5.82) |
| **Creatinine** | 1.16 (1.14, 1.18) | 0.77 (0.75, 0.78) | 0.31 (0.30, 0.31) | 0.34 (0.33, 0.34) | 0.69 (0.66, 0.73) | 0.63 (0.62, 0.64) | 1.15 (1.13, 1.16) | 2.34 (2.24, 2.45) | 5.33 (5.22, 5.43) | 5.29 (5.14, 5.44) | 4.06 (3.98, 4.15) | 5.54 (5.35, 5.73) |
| **Hgb** | 1.31 (1.29, 1.33) | 0.65 (0.64, 0.66) | 0.26 (0.26, 0.27) | 0.35 (0.35, 0.36) | 0.71 (0.67, 0.74) | 0.71 (0.70, 0.72) | 1.14 (1.12, 1.16) | 1.70 (1.64, 1.77) | 4.45 (4.37, 4.54) | 4.62 (4.49, 4.74) | 3.86 (3.79, 3.94) | 5.78 (5.60, 5.97) |
| **WBC** | 1.34 (1.32, 1.36) | 0.65 (0.64, 0.66) | 0.27 (0.26, 0.27) | 0.38 (0.38, 0.39) | 0.70 (0.67, 0.74) | 0.75 (0.74, 0.75) | 1.14 (1.12, 1.16) | 1.74 (1.68, 1.81) | 4.45 (4.36, 4.53) | 4.75 (4.62, 4.87) | 3.76 (3.68, 3.83) | 5.67 (5.50, 5.86) |
| **HDL** | 1.05 (1.03, 1.06) | 0.89 (0.88, 0.91) | 0.36 (0.35, 0.36) | 0.42 (0.41, 0.43) | 0.60 (0.57, 0.63) | 0.65 (0.64, 0.66) | 1.04 (1.02, 1.05) | 2.52 (2.44, 2.60) | 3.50 (3.44, 3.56) | 2.69 (2.63, 2.75) | 2.48 (2.43, 2.52) | 2.28 (2.22, 2.34) |
| **LDL** | 1.08 (1.07, 1.10) | 0.91 (0.89, 0.92) | 0.37 (0.36, 0.37) | 0.43 (0.42, 0.44) | 0.57 (0.55, 0.60) | 0.80 (0.79, 0.81) | 1.04 (1.03, 1.06) | 2.90 (2.80, 2.99) | 3.66 (3.59, 3.72) | 2.58 (2.53, 2.64) | 2.49 (2.44, 2.54) | 2.15 (2.09, 2.20) |
| **Triglycerides** | 1.04 (1.03, 1.06) | 0.91 (0.90, 0.93) | 0.37 (0.36, 0.38) | 0.41 (0.41, 0.42) | 0.61 (0.58, 0.64) | 0.65 (0.64, 0.66) | 1.03 (1.02, 1.05) | 2.53 (2.44, 2.61) | 3.57 (3.50, 3.63) | 2.67 (2.61, 2.73) | 2.48 (2.44, 2.53) | 2.30 (2.24, 2.36) |
| **HbA1c** | 0.97 (0.96, 0.99) | 0.88 (0.86, 0.89) | 0.44 (0.43, 0.45) | 0.60 (0.59, 0.61) | 0.80 (0.76, 0.84) | 0.58 (0.58, 0.59) | 1.28 (1.26, 1.30) | 4.33 (4.20, 4.46) | 3.31 (3.25, 3.37) | 2.03 (1.99, 2.08) | 2.14 (2.10, 2.18) | 2.23 (2.17, 2.28) |
| **CRP** | 1.39 (1.36, 1.42) | 0.83 (0.82, 0.85) | 0.45 (0.43, 0.46) | 0.51 (0.49, 0.52) | 0.77 (0.72, 0.82) | 0.81 (0.80, 0.83) | 1.14 (1.11, 1.16) | 1.27 (1.24, 1.31) | 2.03 (1.98, 2.08) | 1.78 (1.74, 1.82) | 3.19 (3.11, 3.27) | 4.81 (4.67, 4.96) |
| **TSH** | 1.45 (1.43, 1.47) | 0.79 (0.78, 0.80) | 0.38 (0.37, 0.39) | 0.43 (0.42, 0.44) | 0.75 (0.71, 0.78) | 0.70 (0.69, 0.71) | 1.02 (1.00, 1.03) | 1.68 (1.63, 1.73) | 2.84 (2.79, 2.89) | 2.62 (2.56, 2.67) | 2.46 (2.41, 2.50) | 2.48 (2.42, 2.54) |

**Supplementary Table 2 | Yale New Haven Health System: ever-measured logistic model.** Odds ratios (95% CI) by covariate, univariate (unadjusted) and adjusted. Because race/ethnicity and neighborhood income are collinear, the adjusted estimates come from two models: the race model (Black and Hispanic indicators; income excluded) provides the adjusted estimates for race, ethnicity and all other covariates, whereas a separate income model (below-median income indicator; race/ethnicity excluded) provides the adjusted estimate for income. CCI, modified Charlson Comorbidity Index (excludes diabetes and cancer). Reference groups: male, age <40, White, non-Hispanic, at/above-median income, BMI <30, absence of each condition.

| **Biomarker** | **Female** | **Age 40-60** | **Age≥60** | **Black** | **Hispanic** | **Low income** | **Obese** | **Diabetes** | **Hypertension** | **Cancer** | **CCI 1-2** | **CCI ≥3** |
|---|---|---|---|---|---|---|---|---|---|---|---|---|
| **Univariate (unadjusted)** | | | | | | | | | | | | |
| **Glucose** | 0.94 (0.92, 0.96) | 1.22 (1.20, 1.24) | 1.39 (1.36, 1.42) | 1.11 (1.08, 1.14) | 1.32 (1.27, 1.36) | 1.21 (1.19, 1.23) | 1.71 (1.68, 1.74) | 3.15 (3.07, 3.23) | 2.90 (2.85, 2.95) | 2.67 (2.61, 2.73) | 1.42 (1.39, 1.45) | 4.81 (4.68, 4.95) |
| **Creatinine** | 0.95 (0.93, 0.96) | 1.22 (1.20, 1.24) | 1.42 (1.39, 1.45) | 1.10 (1.07, 1.12) | 1.30 (1.26, 1.35) | 1.20 (1.18, 1.22) | 1.73 (1.70, 1.76) | 3.15 (3.07, 3.23) | 2.97 (2.92, 3.02) | 2.68 (2.62, 2.74) | 1.45 (1.42, 1.47) | 4.90 (4.76, 5.04) |
| **Hgb** | 1.26 (1.24, 1.28) | 0.99 (0.97, 1.01) | 1.15 (1.13, 1.18) | 1.12 (1.09, 1.14) | 1.39 (1.35, 1.44) | 1.20 (1.18, 1.22) | 1.67 (1.64, 1.70) | 2.33 (2.27, 2.39) | 2.17 (2.13, 2.20) | 2.43 (2.38, 2.48) | 1.37 (1.34, 1.39) | 4.10 (3.99, 4.21) |
| **WBC** | 1.26 (1.24, 1.28) | 0.99 (0.97, 1.01) | 1.15 (1.13, 1.17) | 1.12 (1.09, 1.15) | 1.39 (1.35, 1.44) | 1.20 (1.18, 1.21) | 1.67 (1.64, 1.70) | 2.32 (2.27, 2.38) | 2.17 (2.13, 2.20) | 2.42 (2.37, 2.47) | 1.36 (1.33, 1.38) | 4.09 (3.98, 4.21) |
| **HDL** | 0.78 (0.76, 0.79) | 1.44 (1.41, 1.47) | 1.06 (1.04, 1.09) | 1.19 (1.16, 1.23) | 1.49 (1.44, 1.54) | 1.19 (1.17, 1.21) | 1.70 (1.67, 1.73) | 2.96 (2.89, 3.03) | 2.69 (2.64, 2.75) | 1.35 (1.32, 1.39) | 1.36 (1.33, 1.39) | 2.34 (2.28, 2.40) |
| **LDL** | 0.78 (0.76, 0.79) | 1.44 (1.41, 1.47) | 1.07 (1.04, 1.09) | 1.20 (1.17, 1.24) | 1.51 (1.46, 1.57) | 1.19 (1.17, 1.21) | 1.70 (1.67, 1.74) | 2.98 (2.91, 3.05) | 2.70 (2.64, 2.75) | 1.35 (1.31, 1.38) | 1.36 (1.33, 1.39) | 2.34 (2.28, 2.41) |
| **Triglycerides** | 0.77 (0.76, 0.79) | 1.44 (1.41, 1.47) | 1.08 (1.05, 1.10) | 1.19 (1.16, 1.22) | 1.48 (1.43, 1.54) | 1.19 (1.17, 1.21) | 1.70 (1.66, 1.73) | 3.00 (2.93, 3.08) | 2.71 (2.66, 2.77) | 1.41 (1.37, 1.44) | 1.35 (1.32, 1.38) | 2.46 (2.39, 2.52) |
| **HbA1c** | 0.93 (0.91, 0.95) | 1.34 (1.32, 1.37) | 1.08 (1.05, 1.11) | 1.60 (1.56, 1.65) | 1.97 (1.90, 2.04) | 1.40 (1.37, 1.42) | 2.39 (2.33, 2.44) | 6.36 (6.20, 6.53) | 2.92 (2.86, 2.98) | 1.39 (1.36, 1.43) | 1.36 (1.33, 1.39) | 2.60 (2.53, 2.67) |
| **CRP** | 1.09 (1.05, 1.12) | 1.25 (1.20, 1.29) | 1.19 (1.15, 1.24) | 0.87 (0.83, 0.92) | 1.16 (1.09, 1.23) | 1.07 (1.04, 1.10) | 1.37 (1.33, 1.42) | 2.15 (2.07, 2.24) | 2.01 (1.94, 2.08) | 2.02 (1.95, 2.10) | 1.53 (1.48, 1.58) | 3.42 (3.29, 3.54) |
| **TSH** | 1.36 (1.34, 1.39) | 1.04 (1.02, 1.06) | 0.96 (0.94, 0.98) | 1.04 (1.01, 1.08) | 1.46 (1.41, 1.51) | 1.17 (1.15, 1.19) | 1.51 (1.48, 1.54) | 2.15 (2.09, 2.20) | 1.86 (1.82, 1.90) | 1.65 (1.61, 1.69) | 1.22 (1.20, 1.25) | 2.47 (2.41, 2.54) |
| **Adjusted (mutually adjusted)** | | | | | | | | | | | | |
| **Glucose** | 1.07 (1.05, 1.09) | 0.88 (0.86, 0.90) | 0.56 (0.54, 0.57) | 0.98 (0.96, 1.01) | 1.46 (1.41, 1.51) | 1.17 (1.15, 1.19) | 1.31 (1.29, 1.34) | 1.73 (1.69, 1.79) | 2.09 (2.04, 2.13) | 2.09 (2.03, 2.14) | 1.67 (1.64, 1.71) | 3.70 (3.57, 3.82) |
| **Creatinine** | 1.08 (1.06, 1.10) | 0.88 (0.86, 0.90) | 0.57 (0.55, 0.59) | 0.97 (0.94, 0.99) | 1.45 (1.40, 1.50) | 1.16 (1.14, 1.18) | 1.32 (1.30, 1.35) | 1.71 (1.66, 1.76) | 2.14 (2.09, 2.18) | 2.07 (2.02, 2.12) | 1.70 (1.66, 1.73) | 3.74 (3.62, 3.87) |
| **Hgb** | 1.40 (1.38, 1.43) | 0.66 (0.65, 0.68) | 0.43 (0.41, 0.44) | 0.97 (0.94, 0.99) | 1.43 (1.38, 1.48) | 1.15 (1.13, 1.16) | 1.35 (1.32, 1.38) | 1.44 (1.40, 1.48) | 1.79 (1.76, 1.83) | 2.15 (2.10, 2.21) | 1.68 (1.65, 1.72) | 3.86 (3.73, 3.99) |
| **WBC** | 1.41 (1.38, 1.43) | 0.66 (0.65, 0.68) | 0.42 (0.41, 0.44) | 0.97 (0.94, 1.00) | 1.42 (1.37, 1.48) | 1.14 (1.12, 1.16) | 1.35 (1.32, 1.37) | 1.44 (1.40, 1.48) | 1.80 (1.76, 1.84) | 2.14 (2.09, 2.20) | 1.67 (1.64, 1.71) | 3.85 (3.73, 3.98) |
| **HDL** | 0.87 (0.85, 0.89) | 1.07 (1.04, 1.09) | 0.64 (0.62, 0.66) | 1.01 (0.98, 1.04) | 1.56 (1.50, 1.62) | 1.13 (1.11, 1.15) | 1.24 (1.21, 1.27) | 1.85 (1.80, 1.90) | 2.10 (2.05, 2.15) | 1.06 (1.03, 1.09) | 1.43 (1.40, 1.46) | 1.91 (1.85, 1.97) |
| **LDL** | 0.87 (0.86, 0.89) | 1.07 (1.04, 1.10) | 0.64 (0.62, 0.66) | 1.02 (0.99, 1.05) | 1.58 (1.53, 1.65) | 1.13 (1.11, 1.15) | 1.24 (1.21, 1.27) | 1.86 (1.81, 1.91) | 2.11 (2.05, 2.16) | 1.06 (1.03, 1.09) | 1.43 (1.40, 1.47) | 1.91 (1.85, 1.98) |
| **Triglycerides** | 0.87 (0.85, 0.89) | 1.06 (1.04, 1.09) | 0.63 (0.61, 0.65) | 1.01 (0.98, 1.04) | 1.55 (1.50, 1.61) | 1.13 (1.11, 1.15) | 1.24 (1.21, 1.26) | 1.86 (1.81, 1.91) | 2.10 (2.05, 2.15) | 1.10 (1.07, 1.13) | 1.44 (1.40, 1.47) | 2.00 (1.94, 2.07) |
| **HbA1c** | 1.08 (1.05, 1.10) | 0.92 (0.89, 0.95) | 0.56 (0.54, 0.58) | 1.27 (1.23, 1.31) | 2.03 (1.95, 2.11) | 1.31 (1.28, 1.33) | 1.59 (1.55, 1.63) | 4.27 (4.15, 4.39) | 1.92 (1.87, 1.98) | 1.15 (1.12, 1.19) | 1.40 (1.37, 1.44) | 1.81 (1.74, 1.87) |
| **CRP** | 1.20 (1.16, 1.25) | 0.93 (0.89, 0.97) | 0.58 (0.54, 0.61) | 0.78 (0.74, 0.82) | 1.20 (1.12, 1.28) | 1.02 (0.99, 1.05) | 1.09 (1.05, 1.13) | 1.34 (1.28, 1.40) | 1.39 (1.34, 1.45) | 1.43 (1.37, 1.49) | 2.34 (2.24, 2.43) | 4.21 (4.00, 4.42) |
| **TSH** | 1.49 (1.46, 1.52) | 0.72 (0.70, 0.73) | 0.44 (0.43, 0.46) | 0.88 (0.85, 0.91) | 1.40 (1.35, 1.46) | 1.11 (1.09, 1.13) | 1.20 (1.18, 1.23) | 1.57 (1.52, 1.62) | 1.69 (1.65, 1.74) | 1.48 (1.44, 1.52) | 1.42 (1.39, 1.46) | 2.41 (2.33, 2.49) |

**Supplementary Table 3 | Michigan Genomics Initiative: ever-measured logistic model.** Odds ratios (95% CI) by covariate, univariate (unadjusted) and adjusted. Because race/ethnicity and neighborhood income are collinear, the adjusted estimates come from two models: the race model (Black and Hispanic indicators; income excluded) provides the adjusted estimates for race, ethnicity and all other covariates, whereas a separate income model (below-median income indicator; race/ethnicity excluded) provides the adjusted estimate for income. CCI, modified Charlson Comorbidity Index (excludes diabetes and cancer). Reference groups: male, age <40, White, non-Hispanic, at/above-median income, BMI <30, absence of each condition.

| Biomarker | Female | Age 40-60 | Age≥60 | Black | Hispanic | Low income | Obese | Diabetes | Hypertension | Cancer | CCI 1-2 | CCI≥3 |
|---|---|---|---|---|---|---|---|---|---|---|---|---|
| **Univariate (unadjusted)** | | | | | | | | | | | | |
| **Glucose** | 1.05 (1.01, 1.10) | 1.50 (1.44, 1.57) | 0.80 (0.76, 0.83) | 1.30 (1.19, 1.41) | 0.79 (0.66, 0.93) | 0.56 (0.54, 0.58) | 1.85 (1.78, 1.93) | 3.39 (3.19, 3.59) | 2.85 (2.74, 2.97) | 1.54 (1.48, 1.60) | 1.23 (1.17, 1.29) | 3.86 (3.68, 4.05) |
| **Creatinine** | 1.06 (1.02, 1.10) | 1.49 (1.43, 1.56) | 0.79 (0.76, 0.83) | 1.27 (1.17, 1.38) | 0.79 (0.67, 0.94) | 0.57 (0.55, 0.58) | 1.82 (1.75, 1.89) | 2.96 (2.79, 3.13) | 2.77 (2.65, 2.88) | 1.55 (1.48, 1.61) | 1.23 (1.17, 1.29) | 3.87 (3.69, 4.05) |
| **Hgb** | 1.14 (1.10, 1.19) | 1.41 (1.36, 1.47) | 0.76 (0.73, 0.79) | 1.29 (1.19, 1.40) | 0.81 (0.69, 0.97) | 0.57 (0.55, 0.59) | 1.72 (1.65, 1.79) | 2.49 (2.36, 2.63) | 2.45 (2.35, 2.55) | 1.52 (1.46, 1.58) | 1.22 (1.16, 1.28) | 3.76 (3.59, 3.94) |
| **WBC** | 1.14 (1.10, 1.19) | 1.41 (1.36, 1.47) | 0.76 (0.73, 0.79) | 1.29 (1.18, 1.40) | 0.82 (0.69, 0.98) | 0.57 (0.55, 0.59) | 1.72 (1.65, 1.79) | 2.50 (2.37, 2.64) | 2.45 (2.35, 2.55) | 1.52 (1.46, 1.58) | 1.22 (1.17, 1.28) | 3.77 (3.60, 3.95) |
| **HDL** | 1.11 (1.07, 1.16) | 1.53 (1.47, 1.59) | 0.47 (0.45, 0.50) | 1.88 (1.74, 2.04) | 1.00 (0.83, 1.20) | 0.26 (0.25, 0.27) | 2.08 (1.99, 2.18) | 3.73 (3.55, 3.91) | 3.08 (2.95, 3.22) | 1.07 (1.02, 1.11) | 1.17 (1.11, 1.22) | 2.78 (2.66, 2.90) |
| **LDL** | 1.11 (1.06, 1.16) | 1.54 (1.47, 1.60) | 0.47 (0.45, 0.50) | 1.90 (1.75, 2.05) | 0.99 (0.82, 1.19) | 0.26 (0.25, 0.27) | 2.09 (2.00, 2.19) | 3.78 (3.60, 3.96) | 3.12 (2.99, 3.27) | 1.05 (1.01, 1.10) | 1.17 (1.12, 1.23) | 2.75 (2.63, 2.87) |
| **Triglycerides** | 1.10 (1.05, 1.15) | 1.54 (1.47, 1.60) | 0.48 (0.46, 0.51) | 1.87 (1.73, 2.02) | 0.97 (0.81, 1.17) | 0.27 (0.26, 0.27) | 2.07 (1.98, 2.17) | 3.74 (3.56, 3.92) | 3.10 (2.97, 3.25) | 1.09 (1.04, 1.13) | 1.16 (1.11, 1.22) | 2.82 (2.70, 2.94) |
| **HbA1c** | 1.14 (1.09, 1.19) | 1.48 (1.41, 1.54) | 0.54 (0.51, 0.57) | 2.16 (2.00, 2.34) | 1.07 (0.89, 1.29) | 0.34 (0.33, 0.36) | 2.71 (2.58, 2.84) | 5.93 (5.65, 6.23) | 3.40 (3.24, 3.56) | 1.02 (0.97, 1.06) | 1.17 (1.11, 1.23) | 2.66 (2.55, 2.78) |
| **CRP** | 1.36 (1.29, 1.43) | 1.47 (1.39, 1.55) | 0.52 (0.49, 0.56) | 1.46 (1.33, 1.61) | 0.86 (0.67, 1.10) | 0.51 (0.49, 0.53) | 2.16 (2.04, 2.29) | 2.51 (2.37, 2.66) | 2.53 (2.39, 2.67) | 1.05 (0.99, 1.10) | 1.15 (1.09, 1.22) | 2.75 (2.61, 2.91) |
| **TSH** | 1.60 (1.54, 1.67) | 1.33 (1.28, 1.39) | 0.49 (0.47, 0.52) | 1.56 (1.44, 1.68) | 0.96 (0.81, 1.15) | 0.36 (0.35, 0.37) | 1.84 (1.76, 1.92) | 2.81 (2.68, 2.94) | 2.25 (2.16, 2.35) | 1.36 (1.30, 1.41) | 1.08 (1.03, 1.13) | 3.04 (2.92, 3.17) |
| **Adjusted (mutually adjusted)** | | | | | | | | | | | | |
| **Glucose** | 1.12 (1.07, 1.17) | 0.91 (0.87, 0.96) | 0.42 (0.39, 0.45) | 1.07 (0.98, 1.17) | 0.92 (0.76, 1.11) | 0.54 (0.52, 0.56) | 1.31 (1.25, 1.37) | 1.94 (1.82, 2.08) | 2.02 (1.92, 2.13) | 0.77 (0.73, 0.82) | 2.23 (2.11, 2.35) | 5.66 (5.31, 6.04) |
| **Creatinine** | 1.12 (1.08, 1.18) | 0.91 (0.86, 0.96) | 0.42 (0.39, 0.45) | 1.05 (0.96, 1.15) | 0.93 (0.77, 1.12) | 0.55 (0.53, 0.56) | 1.31 (1.25, 1.37) | 1.67 (1.57, 1.78) | 2.00 (1.90, 2.11) | 0.77 (0.73, 0.82) | 2.27 (2.15, 2.39) | 5.80 (5.44, 6.18) |
| **Hgb** | 1.21 (1.16, 1.26) | 0.83 (0.78, 0.87) | 0.39 (0.37, 0.42) | 1.07 (0.98, 1.17) | 0.92 (0.77, 1.11) | 0.56 (0.54, 0.58) | 1.27 (1.22, 1.33) | 1.45 (1.36, 1.54) | 1.85 (1.75, 1.94) | 0.79 (0.74, 0.83) | 2.31 (2.19, 2.43) | 5.95 (5.58, 6.34) |
| **WBC** | 1.21 (1.15, 1.26) | 0.83 (0.79, 0.87) | 0.40 (0.37, 0.42) | 1.07 (0.98, 1.17) | 0.94 (0.78, 1.12) | 0.57 (0.55, 0.59) | 1.27 (1.22, 1.33) | 1.46 (1.37, 1.55) | 1.85 (1.75, 1.94) | 0.78 (0.74, 0.83) | 2.31 (2.19, 2.44) | 5.98 (5.62, 6.38) |
| **HDL** | 1.14 (1.09, 1.20) | 0.60 (0.57, 0.64) | 0.19 (0.18, 0.20) | 1.36 (1.25, 1.49) | 1.08 (0.88, 1.34) | 0.24 (0.23, 0.25) | 1.28 (1.22, 1.35) | 2.40 (2.27, 2.54) | 2.48 (2.34, 2.63) | 0.59 (0.55, 0.62) | 2.42 (2.27, 2.58) | 5.70 (5.30, 6.13) |
| **LDL** | 1.14 (1.09, 1.19) | 0.61 (0.57, 0.64) | 0.19 (0.18, 0.20) | 1.37 (1.25, 1.49) | 1.07 (0.87, 1.32) | 0.24 (0.23, 0.25) | 1.28 (1.21, 1.34) | 2.43 (2.30, 2.58) | 2.52 (2.38, 2.67) | 0.58 (0.54, 0.61) | 2.40 (2.25, 2.56) | 5.61 (5.22, 6.03) |
| **Triglycerides** | 1.12 (1.07, 1.18) | 0.61 (0.57, 0.64) | 0.19 (0.18, 0.21) | 1.36 (1.24, 1.48) | 1.05 (0.86, 1.30) | 0.24 (0.23, 0.25) | 1.27 (1.21, 1.34) | 2.40 (2.26, 2.54) | 2.49 (2.35, 2.63) | 0.59 (0.56, 0.63) | 2.42 (2.27, 2.59) | 5.74 (5.34, 6.17) |
| **HbA1c** | 1.22 (1.16, 1.28) | 0.61 (0.57, 0.65) | 0.22 (0.20, 0.24) | 1.56 (1.43, 1.71) | 1.20 (0.97, 1.49) | 0.31 (0.30, 0.32) | 1.61 (1.53, 1.70) | 4.09 (3.86, 4.33) | 2.26 (2.12, 2.39) | 0.57 (0.53, 0.61) | 2.23 (2.08, 2.38) | 4.86 (4.51, 5.24) |
| **CRP** | 1.39 (1.32, 1.48) | 0.72 (0.67, 0.77) | 0.30 (0.28, 0.33) | 1.03 (0.93, 1.14) | 0.92 (0.70, 1.19) | 0.54 (0.52, 0.57) | 1.49 (1.40, 1.58) | 1.48 (1.39, 1.58) | 1.86 (1.73, 1.99) | 0.57 (0.53, 0.62) | 2.73 (2.51, 2.98) | 6.20 (5.66, 6.79) |
| **TSH** | 1.75 (1.67, 1.83) | 0.51 (0.48, 0.54) | 0.18 (0.17, 0.20) | 1.14 (1.04, 1.24) | 0.99 (0.81, 1.20) | 0.35 (0.34, 0.37) | 1.27 (1.21, 1.33) | 1.96 (1.85, 2.07) | 1.93 (1.83, 2.04) | 0.85 (0.80, 0.90) | 2.36 (2.22, 2.50) | 5.81 (5.43, 6.21) |

### S2. Per-visit observation models

This section presents per-visit generalized estimating equation models for whether each biomarker was recorded at a given outpatient visit. Supplementary Tables 4-6 report cohort-specific univariate associations. Supplementary Figures 1 and 2 summarize selected adjusted associations across cohorts for sociodemographic characteristics, comorbidity burden and prior measurement of the same biomarker.

**Supplementary Table 4 | All of Us: univariate per-visit observation model.** Unadjusted odds ratios (95% confidence intervals) are shown for biomarker recording at a visit, separately for each biomarker and covariate, from the generalized estimating equation observation model.

| Biomarker | Female | Age 40-60 | Age≥60 | Black | Hispanic | Low income | Obese | Diabetes | Hypertension | Cancer | CCI 1-2 | CCI≥3 | Prior meas. |
|---|---|---|---|---|---|---|---|---|---|---|---|---|---|
| **Glucose** | 0.85 (0.84, 0.86) | 1.12 (1.11, 1.13) | 1.10 (1.09, 1.12) | 0.94 (0.92, 0.95) | 0.93 (0.89, 0.97) | 1.03 (1.02, 1.04) | 1.03 (1.02, 1.04) | 1.31 (1.29, 1.33) | 1.43 (1.42, 1.45) | 1.37 (1.36, 1.39) | 1.04 (1.03, 1.05) | 1.46 (1.44, 1.48) | 1.70 (1.69, 1.72) |
| **Creatinine** | 0.84 (0.83, 0.85) | 1.13 (1.12, 1.14) | 1.12 (1.11, 1.14) | 0.94 (0.93, 0.96) | 0.91 (0.87, 0.94) | 1.02 (1.01, 1.03) | 1.03 (1.02, 1.04) | 1.30 (1.29, 1.32) | 1.46 (1.45, 1.47) | 1.39 (1.37, 1.41) | 1.06 (1.05, 1.07) | 1.47 (1.45, 1.49) | 1.68 (1.66, 1.69) |
| **Hgb** | 0.95 (0.93, 0.96) | 1.01 (1.00, 1.03) | 0.98 (0.96, 0.99) | 0.93 (0.92, 0.95) | 0.97 (0.93, 1.02) | 1.02 (1.01, 1.03) | 0.94 (0.93, 0.95) | 1.00 (0.99, 1.02) | 1.13 (1.12, 1.15) | 1.42 (1.41, 1.44) | 1.05 (1.04, 1.06) | 1.40 (1.38, 1.42) | 2.08 (2.06, 2.10) |
| **WBC** | 0.95 (0.94, 0.96) | 1.01 (1.00, 1.02) | 0.98 (0.97, 0.99) | 0.93 (0.92, 0.95) | 0.98 (0.94, 1.03) | 1.04 (1.03, 1.05) | 0.94 (0.93, 0.95) | 0.98 (0.96, 1.00) | 1.13 (1.12, 1.14) | 1.43 (1.41, 1.45) | 1.05 (1.04, 1.06) | 1.38 (1.36, 1.40) | 2.15 (2.14, 2.17) |
| **HDL** | 0.76 (0.75, 0.76) | 1.29 (1.27, 1.30) | 0.95 (0.94, 0.97) | 0.75 (0.74, 0.76) | 0.83 (0.80, 0.86) | 1.02 (1.01, 1.03) | 1.01 (1.00, 1.02) | 1.03 (1.01, 1.04) | 1.21 (1.20, 1.22) | 0.85 (0.84, 0.86) | 0.92 (0.91, 0.93) | 0.76 (0.75, 0.77) | 0.64 (0.62, 0.65) |
| **LDL** | 0.78 (0.77, 0.79) | 1.28 (1.27, 1.30) | 0.98 (0.97, 0.99) | 0.80 (0.79, 0.81) | 0.81 (0.78, 0.84) | 1.12 (1.11, 1.13) | 1.04 (1.03, 1.05) | 1.04 (1.03, 1.06) | 1.24 (1.23, 1.26) | 0.83 (0.82, 0.84) | 0.92 (0.91, 0.93) | 0.75 (0.74, 0.76) | 0.62 (0.61, 0.63) |
| **Triglycerides** | 0.75 (0.74, 0.76) | 1.27 (1.26, 1.28) | 0.99 (0.98, 1.00) | 0.73 (0.72, 0.74) | 0.85 (0.81, 0.88) | 1.02 (1.01, 1.03) | 1.01 (1.00, 1.02) | 1.04 (1.02, 1.06) | 1.23 (1.21, 1.24) | 0.86 (0.85, 0.87) | 0.93 (0.92, 0.93) | 0.77 (0.76, 0.78) | 0.67 (0.66, 0.69) |
| **HbA1c** | 0.70 (0.69, 0.71) | 1.23 (1.21, 1.25) | 0.98 (0.96, 1.00) | 1.30 (1.27, 1.32) | 1.02 (0.97, 1.07) | 0.94 (0.92, 0.95) | 1.70 (1.68, 1.73) | 3.38 (3.33, 3.43) | 2.09 (2.06, 2.12) | 0.90 (0.89, 0.92) | 1.09 (1.07, 1.10) | 1.18 (1.15, 1.20) | 2.51 (2.47, 2.54) |
| **CRP** | 1.25 (1.21, 1.30) | 0.95 (0.92, 0.98) | 0.85 (0.82, 0.88) | 0.78 (0.75, 0.82) | 1.05 (0.94, 1.17) | 1.00 (0.97, 1.03) | 0.99 (0.96, 1.02) | 0.87 (0.84, 0.91) | 0.98 (0.95, 1.01) | 0.89 (0.86, 0.92) | 1.45 (1.41, 1.49) | 1.24 (1.20, 1.29) | 11.55 (11.18, 11.94) |
| **TSH** | 1.35 (1.33, 1.37) | 1.02 (1.01, 1.04) | 0.91 (0.89, 0.92) | 0.67 (0.66, 0.68) | 1.05 (1.00, 1.09) | 1.00 (0.99, 1.01) | 0.96 (0.95, 0.97) | 0.81 (0.80, 0.83) | 0.94 (0.92, 0.95) | 0.97 (0.96, 0.99) | 0.95 (0.94, 0.96) | 0.71 (0.70, 0.72) | 1.64 (1.61, 1.67) |

**Supplementary Table 5 | Yale New Haven Health System: univariate per-visit observation model.** Unadjusted odds ratios (95% confidence intervals) are shown for biomarker recording at a visit, separately for each biomarker and covariate, from the generalized estimating equation observation model.

| Biomarker | Female | Age 40-60 | Age≥60 | Black | Hispanic | Low income | Obese | Diabetes | Hypertension | Cancer | CCI 1-2 | CCI≥3 | Prior meas. |
|---|---|---|---|---|---|---|---|---|---|---|---|---|---|
| **Glucose** | 0.78 (0.75, 0.80) | 1.22 (1.19, 1.26) | 1.77 (1.72, 1.83) | 0.95 (0.91, 0.99) | 0.76 (0.73, 0.79) | 1.01 (0.98, 1.03) | 1.03 (1.01, 1.06) | 1.70 (1.65, 1.76) | 1.88 (1.84, 1.93) | 3.46 (3.34, 3.57) | 1.02 (1.00, 1.05) | 3.32 (3.21, 3.45) | 2.40 (2.37, 2.44) |
| **Creatinine** | 0.78 (0.76, 0.80) | 1.22 (1.18, 1.25) | 1.79 (1.74, 1.85) | 0.94 (0.90, 0.99) | 0.75 (0.72, 0.79) | 1.00 (0.97, 1.02) | 1.03 (1.01, 1.06) | 1.69 (1.64, 1.75) | 1.91 (1.86, 1.96) | 3.39 (3.28, 3.50) | 1.04 (1.01, 1.07) | 3.31 (3.20, 3.43) | 2.38 (2.34, 2.41) |
| **Hgb** | 0.98 (0.95, 1.01) | 1.01 (0.98, 1.04) | 1.50 (1.44, 1.55) | 0.92 (0.88, 0.97) | 0.79 (0.76, 0.83) | 1.00 (0.98, 1.03) | 0.96 (0.93, 0.98) | 1.22 (1.17, 1.27) | 1.39 (1.35, 1.43) | 3.44 (3.32, 3.56) | 0.96 (0.93, 0.98) | 3.05 (2.94, 3.17) | 2.47 (2.44, 2.51) |
| **WBC** | 0.98 (0.95, 1.01) | 1.01 (0.98, 1.04) | 1.50 (1.45, 1.55) | 0.93 (0.88, 0.97) | 0.79 (0.76, 0.82) | 1.00 (0.97, 1.03) | 0.96 (0.93, 0.98) | 1.22 (1.17, 1.27) | 1.39 (1.35, 1.43) | 3.46 (3.34, 3.58) | 0.95 (0.93, 0.98) | 3.07 (2.95, 3.19) | 2.49 (2.46, 2.52) |
| **HDL** | 0.66 (0.65, 0.68) | 1.45 (1.42, 1.49) | 1.17 (1.14, 1.21) | 1.01 (0.98, 1.04) | 1.03 (0.99, 1.07) | 0.97 (0.95, 0.99) | 1.29 (1.26, 1.31) | 1.86 (1.80, 1.91) | 1.83 (1.79, 1.87) | 0.71 (0.68, 0.73) | 1.11 (1.09, 1.14) | 0.86 (0.83, 0.89) | 0.66 (0.64, 0.69) |
| **LDL** | 0.67 (0.65, 0.68) | 1.45 (1.42, 1.49) | 1.18 (1.14, 1.21) | 1.03 (1.00, 1.06) | 1.06 (1.02, 1.10) | 0.97 (0.95, 0.99) | 1.28 (1.26, 1.31) | 1.88 (1.82, 1.93) | 1.83 (1.79, 1.88) | 0.70 (0.68, 0.73) | 1.12 (1.09, 1.15) | 0.86 (0.83, 0.90) | 0.65 (0.62, 0.67) |
| **Triglycerides** | 0.66 (0.64, 0.67) | 1.45 (1.41, 1.49) | 1.19 (1.15, 1.23) | 1.02 (0.97, 1.08) | 1.01 (0.97, 1.05) | 0.97 (0.95, 1.00) | 1.26 (1.23, 1.29) | 1.88 (1.81, 1.95) | 1.85 (1.80, 1.90) | 0.82 (0.78, 0.87) | 1.12 (1.09, 1.15) | 0.94 (0.88, 1.00) | 0.94 (0.85, 1.04) |
| **HbA1c** | 0.72 (0.70, 0.74) | 1.45 (1.40, 1.49) | 1.16 (1.11, 1.20) | 1.52 (1.47, 1.58) | 1.43 (1.37, 1.50) | 1.20 (1.17, 1.23) | 1.86 (1.81, 1.92) | 7.26 (7.05, 7.48) | 2.40 (2.33, 2.46) | 0.71 (0.68, 0.74) | 1.16 (1.13, 1.19) | 1.15 (1.10, 1.20) | 2.33 (2.27, 2.39) |
| **CRP** | 1.04 (0.98, 1.10) | 1.21 (1.14, 1.28) | 1.17 (1.10, 1.26) | 0.75 (0.68, 0.83) | 0.85 (0.77, 0.94) | 0.87 (0.83, 0.92) | 0.92 (0.87, 0.98) | 1.28 (1.17, 1.39) | 1.16 (1.10, 1.23) | 1.13 (1.05, 1.22) | 1.63 (1.54, 1.72) | 1.55 (1.43, 1.68) | 11.28 (10.59, 12.02) |
| **TSH** | 1.28 (1.24, 1.32) | 1.03 (1.00, 1.06) | 1.05 (1.01, 1.09) | 0.79 (0.76, 0.83) | 0.97 (0.93, 1.02) | 0.92 (0.90, 0.94) | 1.01 (0.98, 1.03) | 1.17 (1.12, 1.21) | 1.13 (1.10, 1.16) | 1.35 (1.29, 1.41) | 0.86 (0.83, 0.88) | 1.56 (1.48, 1.65) | 1.72 (1.65, 1.79) |

**Supplementary Table 6 | Michigan Genomics Initiative: univariate per-visit observation model.** Unadjusted odds ratios (95% confidence intervals) are shown for biomarker recording at a visit, separately for each biomarker and covariate, from the generalized estimating equation observation model.

| Biomarker | Female | Age 40-60 | Age≥60 | Black | Hispanic | Low income | Obese | Diabetes | Hypertension | Cancer | CCI 1-2 | CCI≥3 | Prior meas. |
|---|---|---|---|---|---|---|---|---|---|---|---|---|---|
| **Glucose** | 0.72 (0.70, 0.74) | 1.10 (1.07, 1.13) | 1.38 (1.33, 1.43) | 1.00 (0.95, 1.05) | 0.84 (0.73, 0.98) | 1.40 (1.37, 1.43) | 1.06 (1.03, 1.09) | 2.10 (2.04, 2.16) | 1.79 (1.74, 1.84) | 1.82 (1.77, 1.87) | 0.86 (0.84, 0.88) | 2.28 (2.22, 2.34) | 2.70 (2.67, 2.74) |
| **Creatinine** | 0.75 (0.73, 0.77) | 1.08 (1.05, 1.11) | 1.44 (1.39, 1.49) | 1.00 (0.95, 1.05) | 0.85 (0.73, 0.98) | 1.44 (1.41, 1.47) | 0.99 (0.97, 1.02) | 1.37 (1.33, 1.41) | 1.64 (1.60, 1.68) | 2.02 (1.96, 2.07) | 0.87 (0.85, 0.90) | 2.41 (2.35, 2.47) | 2.76 (2.72, 2.80) |
| **Hgb** | 0.84 (0.82, 0.87) | 0.99 (0.95, 1.02) | 1.31 (1.26, 1.36) | 1.00 (0.94, 1.06) | 0.96 (0.80, 1.15) | 1.52 (1.48, 1.56) | 0.88 (0.85, 0.90) | 1.02 (0.98, 1.05) | 1.25 (1.22, 1.29) | 2.11 (2.04, 2.18) | 0.85 (0.82, 0.87) | 2.38 (2.32, 2.45) | 3.33 (3.28, 3.38) |
| **WBC** | 0.84 (0.81, 0.86) | 0.99 (0.96, 1.02) | 1.31 (1.26, 1.36) | 1.00 (0.95, 1.06) | 0.96 (0.80, 1.15) | 1.54 (1.50, 1.58) | 0.88 (0.85, 0.90) | 1.04 (1.00, 1.07) | 1.27 (1.24, 1.31) | 2.11 (2.04, 2.18) | 0.84 (0.82, 0.87) | 2.42 (2.36, 2.49) | 3.36 (3.31, 3.40) |
| **HDL** | 0.74 (0.71, 0.77) | 1.26 (1.21, 1.30) | 0.95 (0.90, 0.99) | 1.08 (1.02, 1.15) | 1.04 (0.86, 1.25) | 0.73 (0.71, 0.76) | 1.32 (1.27, 1.36) | 2.09 (2.02, 2.16) | 1.85 (1.79, 1.91) | 0.70 (0.68, 0.73) | 1.18 (1.15, 1.22) | 0.92 (0.89, 0.95) | 1.21 (1.14, 1.28) |
| **LDL** | 0.74 (0.71, 0.76) | 1.26 (1.22, 1.31) | 0.94 (0.90, 0.99) | 1.09 (1.02, 1.15) | 1.01 (0.84, 1.22) | 0.73 (0.71, 0.76) | 1.33 (1.28, 1.37) | 2.14 (2.07, 2.22) | 1.88 (1.83, 1.95) | 0.69 (0.67, 0.72) | 1.20 (1.16, 1.24) | 0.91 (0.88, 0.94) | 1.17 (1.11, 1.25) |
| **Triglycerides** | 0.73 (0.71, 0.76) | 1.26 (1.21, 1.31) | 0.95 (0.90, 1.00) | 1.08 (1.02, 1.15) | 1.02 (0.84, 1.22) | 0.74 (0.72, 0.77) | 1.31 (1.27, 1.36) | 2.10 (2.03, 2.17) | 1.87 (1.81, 1.93) | 0.72 (0.69, 0.75) | 1.19 (1.16, 1.23) | 0.93 (0.90, 0.96) | 1.25 (1.17, 1.32) |
| **HbA1c** | 0.75 (0.71, 0.80) | 1.21 (1.14, 1.28) | 0.87 (0.81, 0.94) | 1.42 (1.30, 1.55) | 0.94 (0.71, 1.24) | 0.83 (0.79, 0.87) | 2.06 (1.95, 2.18) | 12.55 (12.11, 13.01) | 2.89 (2.75, 3.04) | 0.68 (0.65, 0.72) | 1.18 (1.12, 1.23) | 1.08 (1.04, 1.13) | 6.55 (6.37, 6.74) |
| **CRP** | 1.13 (1.05, 1.22) | 1.01 (0.94, 1.09) | 0.82 (0.75, 0.90) | 1.13 (1.00, 1.28) | 1.27 (0.79, 2.05) | 1.31 (1.23, 1.39) | 1.19 (1.11, 1.27) | 0.82 (0.76, 0.88) | 1.01 (0.95, 1.08) | 0.64 (0.60, 0.69) | 1.43 (1.34, 1.52) | 0.99 (0.93, 1.06) | 9.65 (9.30, 10.02) |
| **TSH** | 1.44 (1.39, 1.50) | 0.98 (0.95, 1.02) | 0.93 (0.89, 0.98) | 0.94 (0.88, 1.00) | 0.94 (0.80, 1.11) | 0.95 (0.92, 0.98) | 1.12 (1.08, 1.16) | 1.26 (1.22, 1.31) | 1.10 (1.06, 1.14) | 1.14 (1.09, 1.18) | 0.95 (0.92, 0.98) | 1.20 (1.15, 1.24) | 2.40 (2.30, 2.51) |

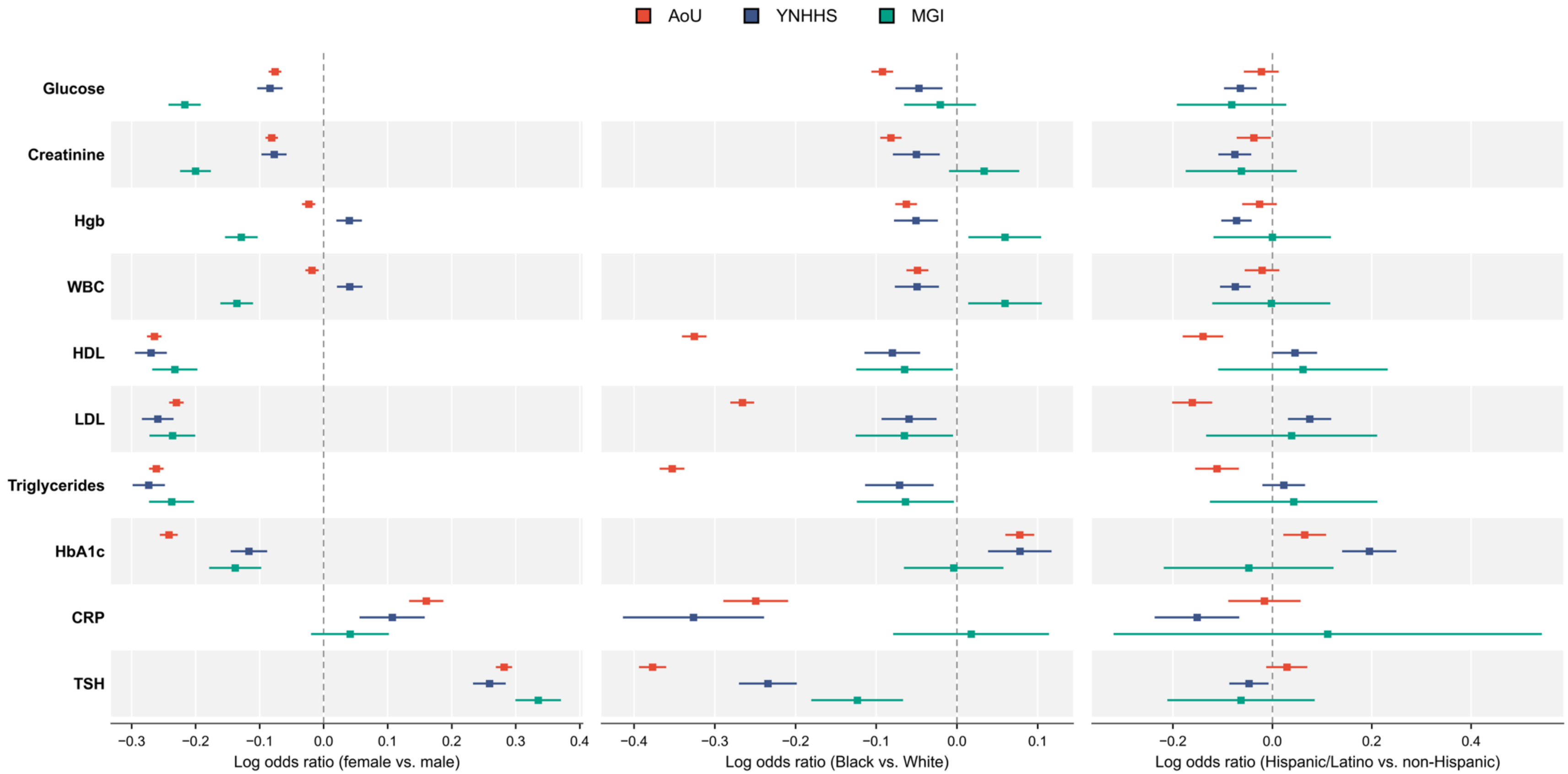


**Supplementary Figure 1 | Sex, race and ethnicity associations with biomarker-specific observation.** Points denote adjusted log-odds ratios and 95% confidence intervals for biomarker recording at an outpatient visit by female versus male sex, Black versus White race and Hispanic/Latino versus non-Hispanic ethnicity, estimated separately within All of Us (AoU; red), Yale New Haven Health System (YNHHS; blue) and the Michigan Genomics Initiative (MGI; green). Dashed vertical lines indicate no association.

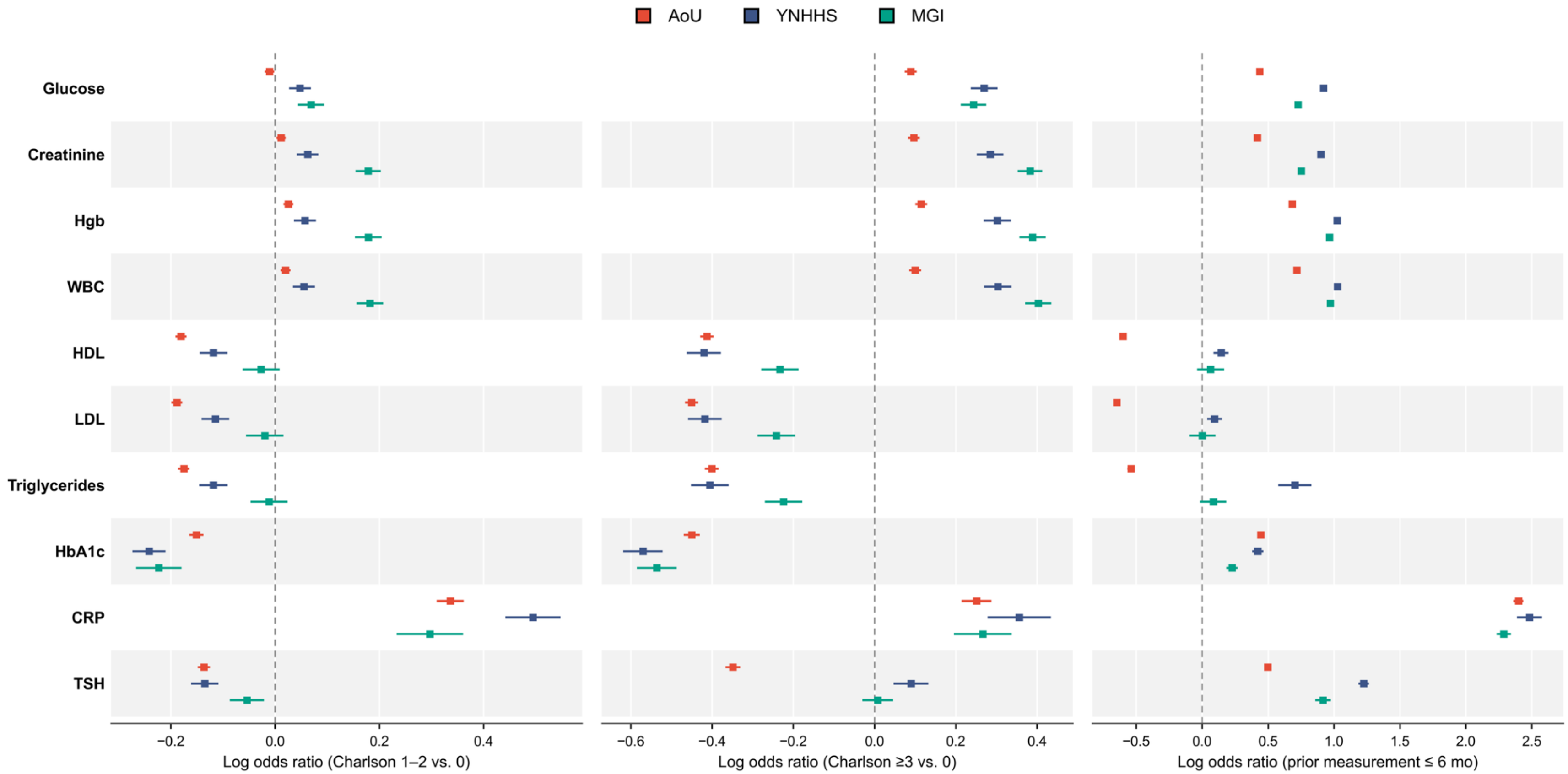


**Supplementary Figure 2 | Comorbidity burden and prior-measurement associations with biomarker-specific observation.** Points denote adjusted log-odds ratios and 95% confidence intervals for recording each biomarker at an outpatient visit by modified Charlson Comorbidity Index category (1–2 and ≥3, each versus 0) and by prior measurement count of the same biomarker within the preceding 6 months. The panels show that a higher comorbidity burden increases recording of some biomarkers while decreasing recording of others in all three cohorts, and that a recent prior measurement increases recording of nearly every biomarker, the exception being the lipid biomarkers in AoU. Cohorts are All of Us (AoU; red), Yale New Haven Health System (YNHHS; blue) and the Michigan Genomics Initiative (MGI; green).

S3. Simulation study

**Simulation design.** We conducted 1,000 Monte Carlo replicates under a two-stage EHR clinical recording process with IP and IO. In each replicate, we generated n = 2,000 participants over a common follow-up window [0, 120]. Baseline covariates were generated as $X_i \sim \text{Bernoulli}(0.5)$ and $Z_i \sim N(0, 1)$, and a shared latent factor $U_i \sim N(0, 1)$ linked the visit, observation and outcome processes. Hence, the shared latent factor $U_i$ induces both IP and IO.

Visits were generated from a homogeneous Poisson process with subject-specific intensity $\lambda_i(t) = \eta_i \exp(-2.8 + 0.5X_i + 0.5Z_i)$ where $\eta_i = \exp(-0.0125 + 0.5U_i)$. Conditional on a visit at time $t$, biomarker recording followed a probit mixed-effects model, $P\{R_i(t) = 1 \mid dN_i(t) = 1, X_i, Z_i, q_{0i}, q_{Z_i}\} = \Phi\{0.15 - 0.45Z_i - 0.25X_i + (-0.25 + 0.25Z_i)U_i + q_{0i} + q_{Z_i}Z_i\}$ where $\Phi(\cdot)$ denotes the standard normal cumulative distribution function, $q_{0i} \sim N(0, 0.25^2)$ and $q_{Z_i} \sim N(0, 0.15^2)$.

The longitudinal biomarker outcome was generated from

$$Y_i(t) = -2 + \beta_Z Z_i + 0.5X_i + b_{1i} + b_{2i}Z_i + \epsilon_i(t), \qquad \epsilon_i(t) \sim N(0, 0.75^2),$$

with outcome random effects linked to the shared latent factor through

$$b_{1i} = 0.45U_i + e_{1i}, \qquad e_{1i} \sim N(0, 0.5^2)$$

and

$$b_{2i} = 0.45U_i + e_{2i}, \qquad e_{2i} \sim N(0, 0.8^2).$$

At generated visit time, $Y_i(t)$ was observed only when $R_i(t) = 1$; otherwise, the biomarker value was treated as missing. The target estimand was the fixed effect of $Z_i$ in the outcome model, $\beta_Z = -0.5$. Simulation results are summarized across Monte Carlo replicates. Bias measures systematic departure from $\beta_Z$, whereas RMSE captures both bias and estimator variability; these summaries are shown in Supplementary Figures 3 and 4, respectively.

**Estimators evaluated.** We compared ten estimators of the longitudinal-outcome regression coefficient $\beta_Z$. The first four are summary-statistic regressions that collapse each participant's observed biomarker trajectory to a single statistic and regress that statistic on the baseline covariates $X_i$ and $Z_i$ by ordinary least squares: Summary-mean uses the per-participant arithmetic mean of observed outcomes, Summary-median the per-participant median, and Summary-min and Summary-max the per-participant minimum and maximum, respectively. These four estimators ignore both the visiting and the conditional observation processes and serve as naive comparators against which adjustment methods are evaluated. The fifth estimator is a linear mixed-effects model[1] (LMM) with a participant-specific random intercept fitted by restricted maximum likelihood to all observed outcomes; this is the standard longitudinal analysis under the assumption that observation times are non-informative conditional on covariates.

Three further estimators perform IP-only adjustment. Inverse intensity rate ratio[2] (IIRR) weighting estimates subject-specific visit intensities from an LWYY proportional-rate model on the visit-process covariates and reweights the outcome regression by the reciprocal of each estimated intensity. The joint model for visit process and longitudinal outcome[3] (JMVL-Liang) links the visiting-process intensity and the longitudinal outcome through a single participant-level random effect. Pairwise likelihood[4] estimates the longitudinal mean structure by maximizing a pairwise likelihood for irregularly observed longitudinal outcomes, allowing dependence between observation times and outcomes without specifying the observation-time process. None of these three methods explicitly models the biomarker-specific observation process after a visit.

The remaining two estimators model both stages of EHR data generation. EHRJoint[5] specifies separate submodels for visit occurrence, biomarker recording and the longitudinal outcome, but links the latent structure only to the visit and outcome components; the biomarker-recording model conditions on observed covariates and is therefore missing at random with respect to the latent health process. Clinically Informative Missingness in Electronic Health Records[6] (CIMEHR) instead links all three components through a shared Gaussian latent variable, allowing unmeasured patient heterogeneity to jointly drive visit intensity, biomarker recording and the longitudinal biomarker trajectory.

**Simulation results.** A simulation confirmed that both stages matter for downstream estimation and that methods able to address them exist. With a known covariate effect ($\beta_Z = -0.5$) and a shared latent health process driving visiting, recording and the outcome, a standard LMM was biased for the true effect (mean estimate -0.56; bias -0.06), and adjusting only the visiting stage did not remove the bias (IIRR -0.64, bias -0.14). Estimators that modeled both stages were close to the truth (EHRJoint -0.49, bias 0.01; CIMEHR -0.50, bias 0.00). Full details and the complete comparison are in Supplementary Figure 3.

Adjusting only the visiting process performed worse than no adjustment at all. Relative bias was 12% for the naive LMM and 28% for IIRR weighting, but fell to 2% for EHRJoint and 0% for CIMEHR (Supplementary Figure 3). The amplification arose because the second-stage selection was informative but ignored: the visit-process estimating equation reconciles an outcome trajectory already selected at two stages using weights or random effects calibrated to absorb only one, so the adjustment is not merely incomplete but misdirected. Joint modeling removed this amplification at a modest cost in variance, with RMSE similar to that of the LMM and lower than that of the IP-only estimators (Supplementary Figure 4).

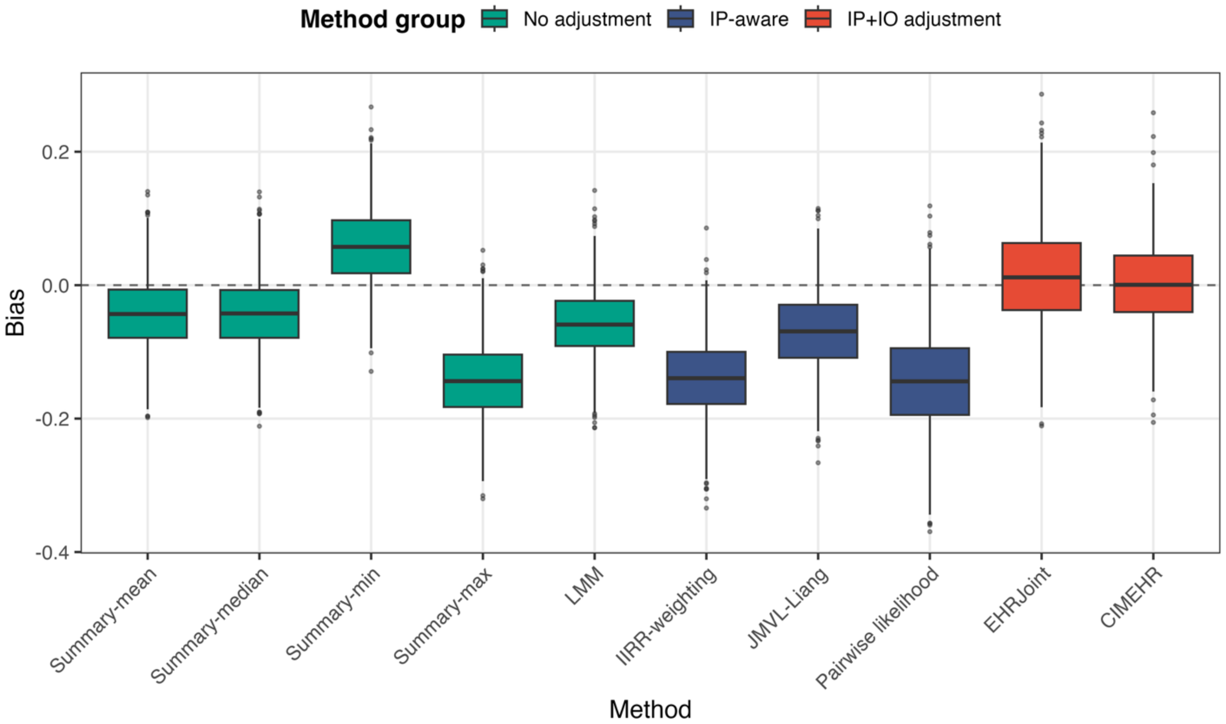


**Supplementary Figure 3 | Bias of estimators in the simulation study.** Box plots show Monte Carlo bias of each estimator of the longitudinal-outcome regression coefficient, computed against the true value of the coefficient, across 1,000 replicates of the joint informative-presence and informative-observation data-generating mechanism described in Methods. Estimators are organized into three classes: (i) naive estimators (green) that ignore both stages of the clinical recording process, comprising Summary-mean, Summary-median, Summary-min and Summary-max (participant-level summary regressions on the corresponding statistic of each participant's observed biomarker trajectory) and a random-intercept linear mixed model (LMM); (ii) IP-only estimators (blue) that adjust for informative visiting but assume biomarker recording is non-informative given the modeled covariates, comprising inverse intensity rate-ratio weighting (IIRR-weighting), the shared-frailty joint visit-outcome model (JMVL-Liang) and pairwise composite likelihood; and (iii) joint IP/IO estimators (red) that model the visiting and conditional observation processes simultaneously, comprising EHRJoint and CIMEHR (Clinically Informative Missingness for EHR data). The dashed horizontal line denotes zero bias.

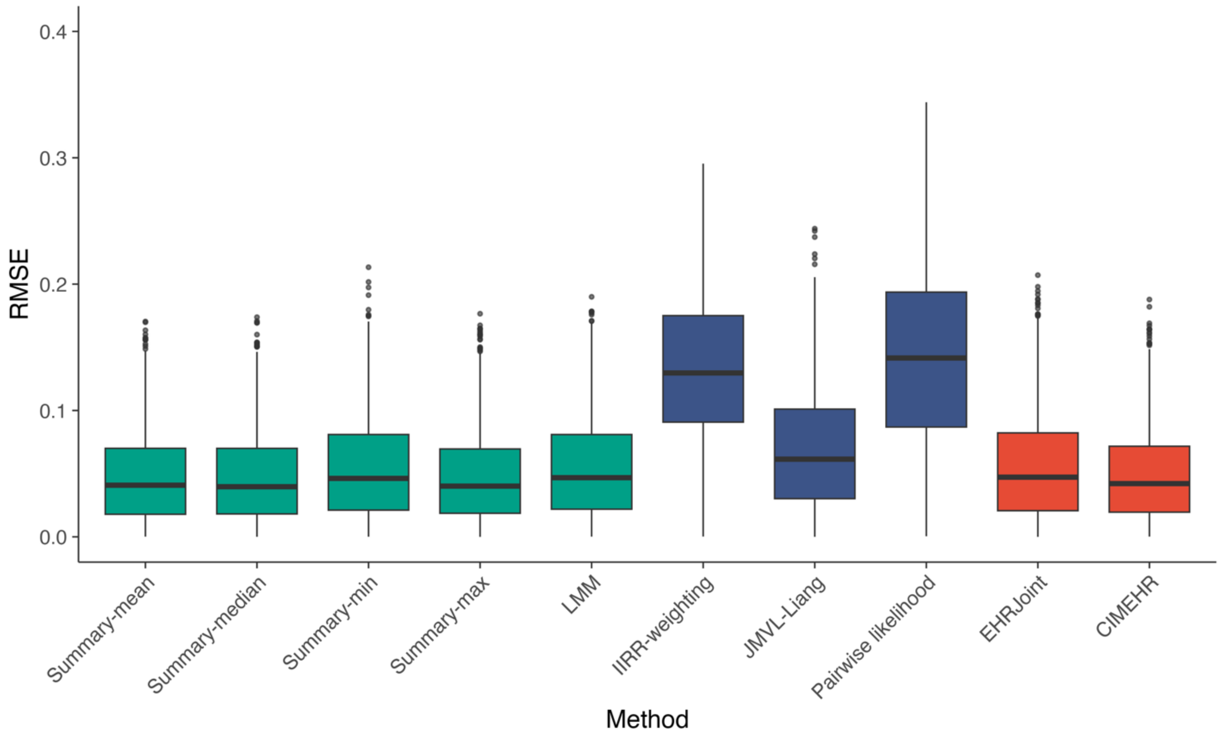


**Supplementary Figure 4 | Root mean squared error (RMSE) of estimators in the simulation study.** Box plots show RMSE relative to the target estimand across 1,000 Monte Carlo replicates for the same estimators as in Supplementary Figure 3. RMSE paralleled the bias results: IIRR-weighting and pairwise likelihood had the highest RMSE, the visit-process joint model was intermediate, and EHRJoint and CIMEHR had lower RMSE with estimates centered closer to the target.

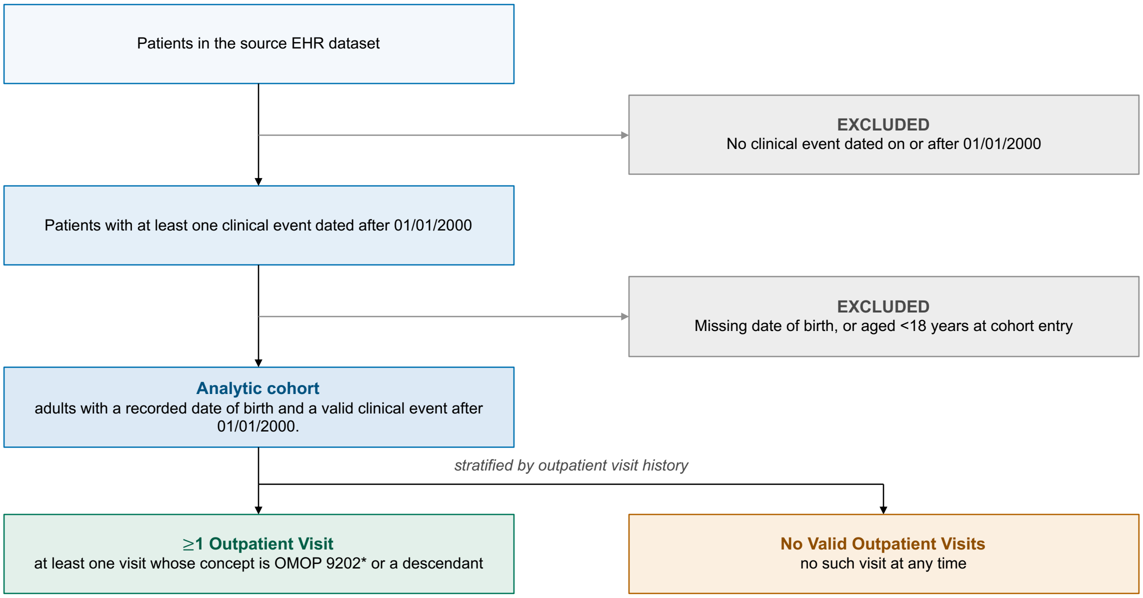


**Supplementary Figure 5 | Definition of the analytic cohort.** Boxes on the left show the criteria applied in sequence to each source electronic health record dataset, and grey boxes on the right the patients removed at each step. Dividing by outpatient visit history removes no patient: both groups (green and orange) remain in the analytic cohort. Outpatient visits were identified from the Observational Medical Outcomes Partnership (OMOP) common data model as visits with concept_id 9202 (Outpatient Visit) or one of its descendants, excluding emergency department and inpatient visits. The same sequence was applied in All of Us, Yale New Haven Health System and the Michigan Genomics Initiative.

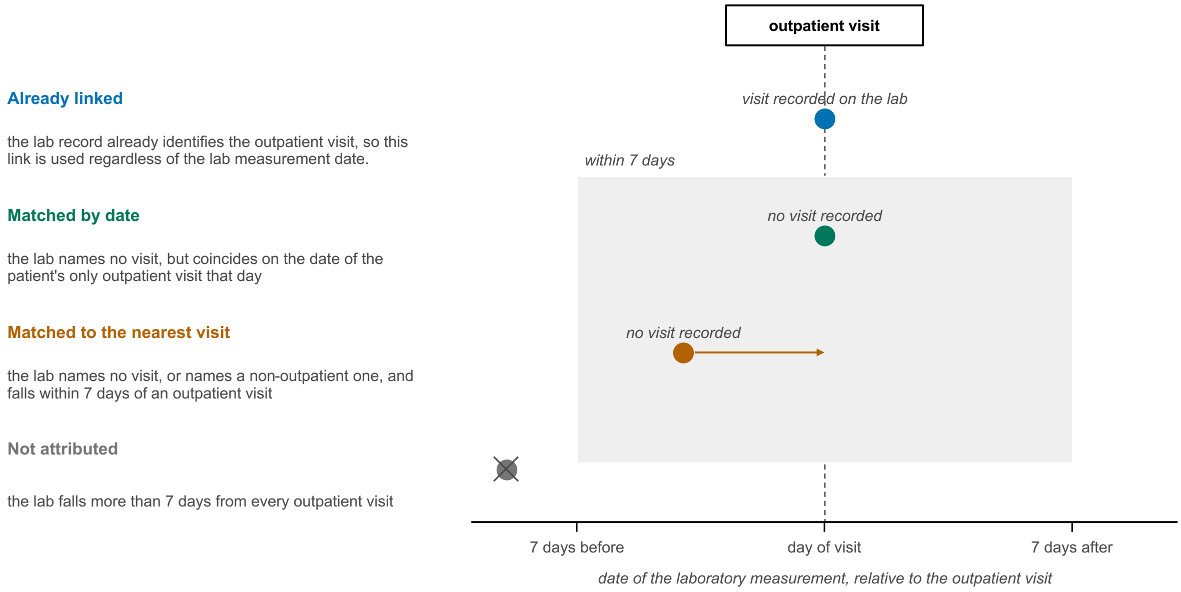


**Supplementary Figure 6 | Mapping of laboratory measurements to outpatient visits.** The left-hand side show the four rules applied in order to each laboratory measurement, each with one example measurement placed on a time axis centered on the outpatient visit; the shaded band spans seven days either side of the visit date. Each measurement is attributed at most once, by the first rule that applies, and only attributed measurements enter the per-visit recording indicator.

### Supplementary References


1. Laird, N. M. & Ware, J. H. Random-effects models for longitudinal data. *Biometrics* **38**, 963–974 (1982).

2. Buzkova, P. & Lumley, T. Longitudinal data analysis for generalized linear models with follow-up dependent on outcome-related variables. *Can. J. Stat.* **35**, 485–500 (2007).

3. Liang, Y., Lu, W. & Ying, Z. Joint modeling and analysis of longitudinal data with informative observation times. *Biometrics* **65**, 377–384 (2009).

4. Chen, Y., Ning, J. & Cai, C. Regression analysis of longitudinal data with irregular and informative observation times. *Biostatistics* **16**, 727–739 (2015).

5. Du, J., Shi, X. & Mukherjee, B. A new statistical approach for joint modeling of longitudinal outcomes measured in electronic health records with clinically informative presence and observation processes. Preprint at https://doi.org/10.48550/arXiv.2410.13113 (2025).

6. Yang, C.-H., Shi, X. & Mukherjee, B. Joint Modeling of Longitudinal EHR Data with Shared Random Effects for Informative Visiting and Observation Processes. Preprint at https://doi.org/10.48550/arXiv.2602.15374 (2026).
